# Long COVID undercoding and prediction from releasable electronic health record summaries in All of Us

**DOI:** 10.64898/2026.10.18.26365858

**Authors:** Amir Asiaee, Kaveh Aryan

**Author notes:** Corresponding author: Amir Asiaee.

## Abstract

**Objective:** To estimate Long COVID undercoding in electronic health records (EHRs) against federal survey prevalence, and to evaluate a predictor of later coding that anyone can rebuild from counts a research enclave releases.

**Materials and Methods:** Among 23,790 All of Us adults with COVID-19, we compared coded shares with Household Pulse Survey prevalence. Among 23,300 adults uncoded at day 90, we predicted a first U09.9 code by day 455. We fitted naive Bayes and linear discriminant models outside the enclave from released counts and compared them with an elastic net fitted inside to the same patients and features. Tests were a random 20% of patients (4,680, 50 coded) and 18 held-out EHR sites (5,760, 60 coded).

**Results:** Under survey-comparability assumptions, about 1 in 9 adults who ever had Long COVID carried its code, and the women-to-men coding ratio was 0.79 (95% sensitivity interval, 0.65–0.95). The released models’ areas under the receiver operating characteristic curve (AUROCs) were 0.68 (95% CI, 0.60–0.75) on the patient test and 0.64 (0.55–0.72) at held-out sites, 0.049 (0.001–0.104) above the elastic net. On the patient test, the elastic net had a higher area under the precision-recall curve (difference 0.062, 0.003–0.122). Released probabilities overestimated coding risk at held-out sites.

**Discussion:** Fitting from released counts did not consistently lower discrimination. Coded outcomes inform disease discrimination only under stated coding assumptions.

**Conclusion:** Long COVID is substantially and unevenly undercoded, and a model rebuilt from public counts predicts its later coding with moderate discrimination.

## 1 Background and Significance

Long COVID affects patients whose electronic health records (EHRs) may never contain its diagnosis code. The ICD-10-CM code U09.9 became available on October 1, 2021, but its sensitivity was 0.15 against the highly symptomatic RECOVER research-index phenotype and 10.5% to 14.9% against a WHO-based chart-review definition in two health systems [1, 2]. Code use and the characteristics of diagnosed patients also vary across clinicians, sites, and demographic groups [3–8]. These differences distort estimates of disease burden and shape what prediction models trained on diagnosis codes learn.

EHR studies have modeled recorded Long COVID and post-COVID conditions using information before and after infection [9–12]. The National COVID Cohort Collaborative (N3C) model reached an area under the receiver operating characteristic curve (AUROC) of 0.72 when transferred to the All of Us Research Program [13], and another All of Us study reported 0.736 [14]. These studies show that recorded Long COVID has a predictable EHR signature. What such a model means depends on when its predictors are measured, which event defines the outcome, and how clinical disease becomes documented. Positive-unlabeled learning makes incomplete labeling explicit, but connecting observed labels to disease still requires assumptions about how codes are assigned [15–17].

A second obstacle arises when the data used to develop a model must stay inside a research enclave. All of Us does not allow models fitted to participant records to leave its Researcher Workbench, but it allows sharing of models fitted outside from dissemination-compliant summaries [18]. Distributed regression and related methods have long supported analysis without pooling patient records [19–22]. A fixed public set of counts offers a simpler route: anyone can rebuild the model without enclave access or a continuing link to the contributing sites. The practical question is how much predictive information survives the feature restrictions and count processing that such a release requires.

Federal surveys and enclave EHR data address complementary parts of this problem. The Household Pulse Survey (HPS) measures self-reported Long COVID prevalence among adults who have had COVID-19 [23]. Comparing that prevalence with EHR coding quantifies the documentation gap under explicit population-comparability assumptions. Within the EHR cohort, matched prediction experiments can then show what each release restriction costs a model that others can rebuild and use.

## 2 Objective

In this paper, we make three contributions. First, we estimate Long COVID coding fractions by sex, age, and race and ethnicity against federal survey prevalence. Second, we evaluate a predictor of a later U09.9 code that anyone can rebuild from released counts. We compare it, on the same patients, with models that must remain in the enclave and separate the effects of feature representation, estimator, feature exclusion, and count processing. Third, we state the assumptions under which coded-outcome evaluation also ranks models by their discrimination of the underlying disease.

## 3 Materials and Methods

### 3.1 Data sources and study population

Reporting follows TRIPOD+AI [24]. We used the All of Us Research Program Registered Tier Dataset v9 (curated data repository R2025Q4R6, cutoff January 1, 2025) [25], in which dates are shifted backward by 1 to 365 days per participant, preserving within-person intervals. HPS estimates came from the Centers for Disease Control and Prevention and National Center for Health Statistics release of the Census Bureau survey. We used 27 periods from June 2022 through September 2024, each reporting the share of adults with prior COVID-19 who ever had or currently had Long COVID [26].

The index was the first U07.1 code on or after shifted date July 2, 2021, so that day 91 always falls after U09.9 became available. We required age 18 years or older at the index, no Long COVID code on or before it, and an EHR observation period spanning days *−*365 to 455. For prediction, we additionally excluded patients with U09.9 or a qualifying SNOMED CT post-COVID code by day 90. The outcome was a first U09.9 code during days 91–455 (Figure 1). We assigned each patient to a site, the EHR source identifier of the index event: a health care organization contributing EHR data for 93.4% of the day-90 cohort and a participant-mediated EHR source for 6.6% (Supplementary Section S9).

**Figure 1:**
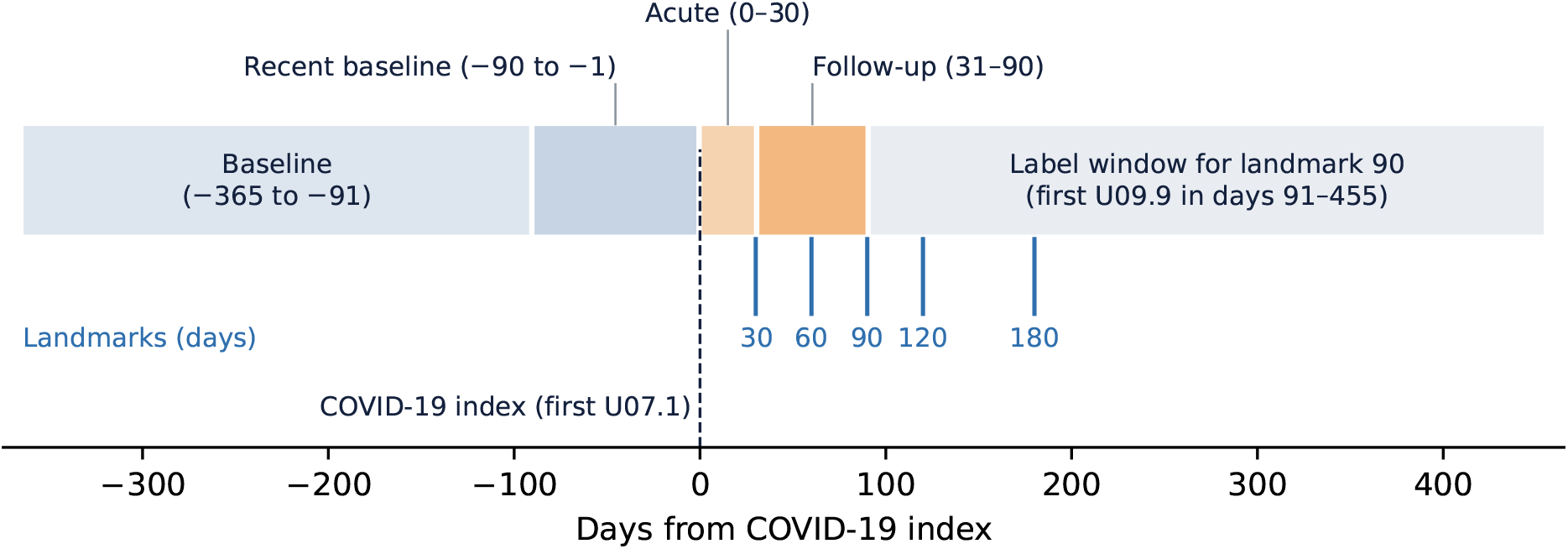
Study design. Predictor windows relative to the COVID-19 index date and the outcome window for the primary day-90 landmark. Blue ticks mark the additional landmarks in Supplementary Section S10. At landmark *L*, the late predictor window runs from day 31 to day *L*, and the outcome window from day *L* + 1 to day 455. Timeline of the predictor windows before and after the COVID-19 index date and of the outcome window, in which a first U09.9 code is ascertained from day 91 to day 455.

### 3.2 Coding fraction and prediction target

For demographic group *g*, let *p_g_* be the coded share during days 1–455 and *π*^HPS^ the survey prevalence. If codes identify survey-defined cases and the cohort and survey prevalences agree, the coding fraction is *c_g_* = *p_g_/π*^HPS^. More generally, with *v_g_* the positive predictive value of the code for the survey-defined condition and *κ_g_* the cohort-to-survey prevalence ratio,

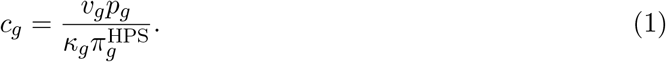

The primary analysis sets *v_g_* = *κ_g_* = 1. We estimated coding fractions and between-group ratios separately for the ever and current survey definitions. Central 95% sensitivity intervals came from 5,000 replicates, each drawing one survey period shared across groups and a binomial coded share. Supplementary Section S2 varies code validity, prevalence comparability, within-period survey error, demographic composition, and age-group weights.

The prediction target is distinct. Among patients eligible at day 90, let *X* contain information available by that day, *C* indicate the later code, and *Y* denote underlying Long COVID during follow-up. Models estimate *q*(*x*) = *P* (*C* = 1 *| X* = *x*) or a score that ranks it, and we evaluate them against the observed *C*. Supplementary Section S5 shows how coded-outcome AUROC relates to AUROC against *Y* when coding errors do not depend on features within disease classes [27]. The survey supplies the population comparison in Equation (1), not individual outcomes for the prediction task.

### 3.3 Features and models

We represented each patient’s chart in two ways. The *binary* representation contained 102 indicators of demographics, prior conditions, acute severity, health care use, symptoms, tests, and medications. Each is 1 when the patient is in the group or has a qualifying record in its window (days *−*365 to *−*91, *−*90 to *−*1, 0–30, or 31–90) and 0 otherwise (Supplementary Table S1). The *rich* representation contained 123 features that keep more of the same information: age and its square, sex, the number of symptom groups coded, and log-transformed counts, such as visits by class and window and distinct dates with each condition group, test, laboratory order, and prescription (Supplementary Section S8). It therefore mixes continuous, count, and binary features. In both, ICD-10-CM predictors came from the condition and observation tables, and other features from CPT, LOINC, and RxNorm records.

Inside the enclave, we fitted elastic-net logistic regression to each representation, which we call the binary and rich elastic nets, with standardized predictors, an *ℓ*_1_ mixing proportion of 0.5, and inverse penalty 0.05. Outside the enclave, we fitted naive Bayes (NB) and linear discriminant analysis (LDA) to the binary representation. These two models can leave the enclave because their fits depend on the data only through counts: NB needs the class counts and each feature’s counts within each class, and LDA also needs the counts of each pair of features occurring together. Released counts therefore suffice to fit them, whereas an elastic net needs every patient’s feature values and must stay inside. Each used empirical-Bayes shrinkage of its feature signals, and LDA also shrank the within-class covariance toward its diagonal with *λ ∈ {*0.1, 0.3, 0.6*}*, giving four candidate specifications in all. Both yield a linear score *r*(*x*) = *β*_0_ + *w*^T^*x*. Supplementary Section S5 derives these fits from counts.

### 3.4 Summary release and matched evaluation

Figure 2 traces the procedure from the cohort to the released artifact. Our release procedure retained a binary feature when all four cells of its feature-by-outcome table exceeded 20. It released those tables, feature margins, and pair counts for retained features, suppressing participant counts of 1–20, rounding other positive counts up to the next multiple of 10 so that a small count cannot be recovered exactly by subtracting released counts, and suppressing a further cell wherever a hidden cell could be recovered from the same table’s totals. A released count *r* therefore stands for a true count between *r −* 9 and *r*. Outside the enclave, we replaced each positive released count with the midpoint of that range, *r −* 4.5, and fitted the models. Unavailable pair moments would use the independence value (Supplementary Section S11).

**Figure 2:**
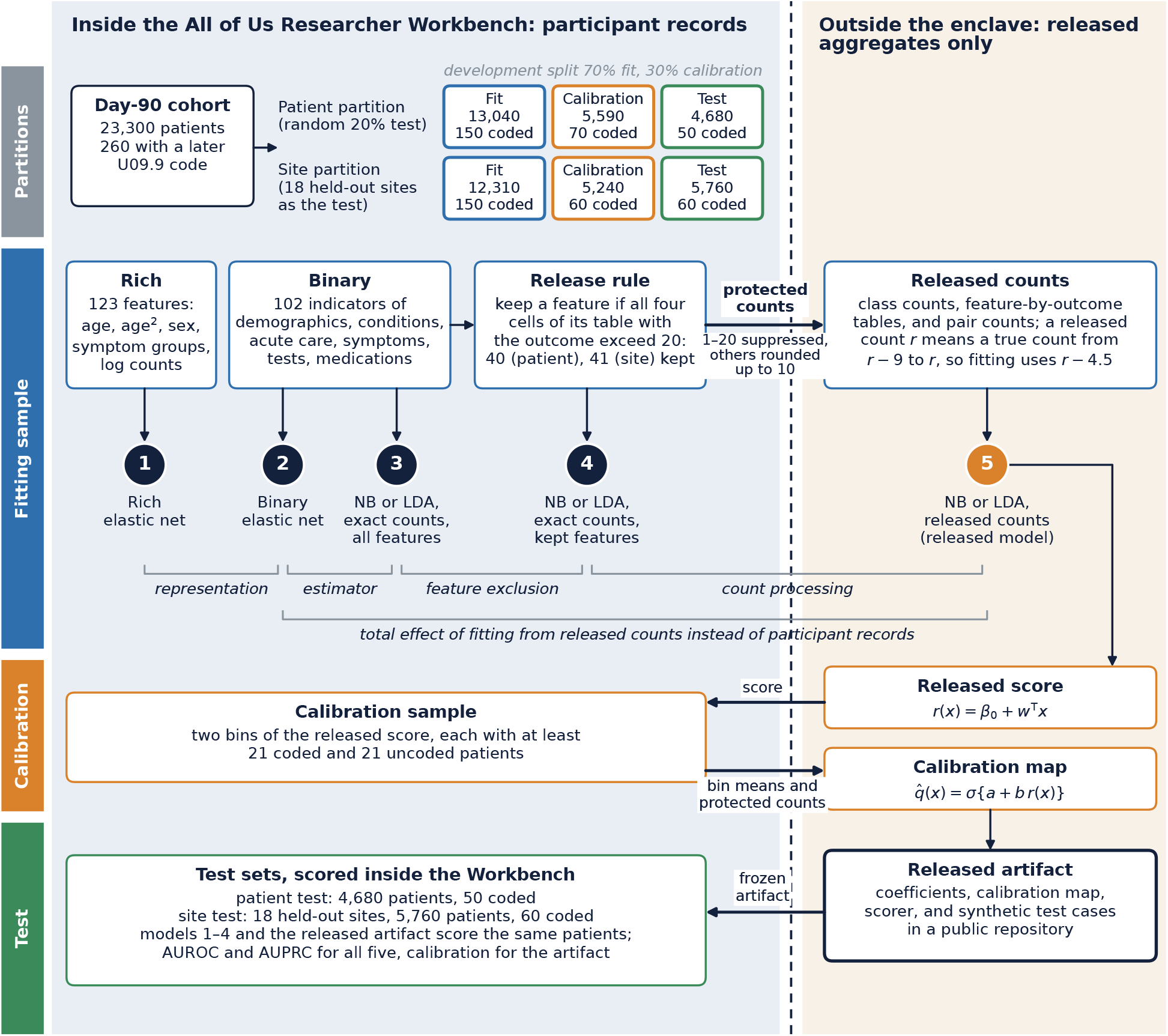
Summary release and matched evaluation. Each partition of the day-90 cohort was divided into fitting, calibration, and test patients, with counts rounded up to the next multiple of 10. All five models were fitted to the same fitting patients: models 1 to 4 inside the enclave and model 5 outside it, from released counts. Models 3 to 5 use the specification that 3-fold cross-validation selected within the fitting sample, repeating every fitting step: NB in the patient partition and LDA with *λ* = 0.6 in the site partition. Brackets mark the paired comparisons in Figure 4. Arrows across the dashed line are the only transfers between the enclave and the outside: protected counts and calibration-bin summaries leave it, and the released score and the frozen artifact return to it. Schematic of two regions, inside the enclave and outside it, separated by a dashed boundary. The day-90 cohort of 23,300 patients is divided into two partitions, each with fitting, calibration, and test patients. In the fitting sample, the rich representation (123 features) and the binary representation (102 indicators) feed five numbered models: two elastic nets and two exact-count NB or LDA models inside, and one NB or LDA model fitted outside from protected counts of the 40 or 41 retained features. The released score returns to form two calibration bins, whose summaries leave to fit the calibration map, and the frozen artifact returns to be scored on the patient and site tests together with models 1 to 4.

We evaluated two outer partitions. The patient test was a random 20% of the day-90 cohort (4,680 patients, 50 with a later code). The site test comprised 18 held-out sites (5,760 patients, 60 with a later code). Like every participant count in this paper, these sizes are rounded up to the next multiple of 10. Within each development partition, 70% of patients formed the fitting sample and 30% the calibration sample.

In each partition we fitted five models to the same fitting patients, numbered as in Figure 2: (1) the elastic net with rich features, (2) the elastic net with binary features, (3) the selected NB or LDA specification from exact counts of all 102 binary features, (4) the same specification from exact counts of the retained features, and (5) the same specification from released counts of the retained features. Model 2 is the participant-level counterpart of the released model, fitted inside the enclave to the same patients and binary features. Comparing models 1 and 2 isolates the representation, 2 and 3 the estimator, 3 and 4 feature exclusion, and 4 and 5 count processing. Comparing models 5 and 2 gives the total effect of fitting from released counts instead of participant records. Three-fold cross-validation within the fitting sample selected the NB or LDA specification by mean AUROC. Each fold repeated feature eligibility, count protection, reconstruction, and fitting, so selection used no calibration or test patient.

After fitting the score outside the enclave, we returned it to the Workbench to form calibration bins from the calibration sample. Each bin held at least 21 coded and 21 uncoded patients, and only its mean score and protected counts were released. Outside the enclave we fitted *q*^(*x*) = *σ{a*+*b r*(*x*)*}*, where *σ* is the logistic function and *b ≥* 0. Each partition yielded two bins, so the two-parameter map reproduces both grouped event rates and leaves no residual for checking fit within the calibration sample. We therefore assessed calibration on the test partitions. The model fitted without the held-out sites is the focal released model, and the model fitted in the patient partition is the matching instance for the patient test.

We report AUROC, area under the precision-recall curve (AUPRC, computed as average precision), and the calibration intercept in the large and calibration slope. Intervals are 95% CIs from 500 participant bootstrap replicates with fixed predictions, stratified by outcome for discrimination and paired across models for differences. Additional cross-validated models, landmark analyses, subgroups, and associations with later symptom codes appear in Supplementary Section S10.

### 3.5 Released artifact and ethics

The released artifact contains feature definitions, coefficients, the calibration map, and hashes of its aggregate inputs and source code, together with a reference scorer and synthetic test cases. The coefficients and calibration map evaluated in the Workbench are the same objects supplied with the release. An existing SMART on FHIR application [28], described in a companion paper [29], runs a separately fitted 57-feature NB model from the same feature registry, which reads FHIR resources instead of the research data model (Supplementary Section S10). All participant records, individual scores, and participant-trained models remained in the Workbench. All of Us participants provide informed consent under the program’s institutional review board protocol, and we analyzed de-identified Registered Tier data as authorized researchers [25].

## 4 Results

### 4.1 Cohort and coding fractions

Of 50,210 adults with a U07.1 code, 23,790 met the cohort criteria and 23,300 remained uncoded at day 90 (Table 1, Supplementary Figure S1). Of these, 260 (1.1%) received a first U09.9 code during days 91–455. The held-out sites had more women and more non-Hispanic White patients than the development sites (Supplementary Table S8).

**Table 1:** Cohort characteristics.

|  |  | Full cohort |  | Landmark cohort (uncoded at day 90) |  |
| --- | --- | --- | --- | --- | --- |
|  |  | Adults, n | Coded in days 1–455, % | Adults, n | Coded in days 91–455, % |
| Sex | Female | 14,870 | 3.2 | 14,550 | 1.2 |
|  | Male | 8,730 | 2.9 | 8,560 | – |
|  | Other/unknown | 200 | – | 200 | – |
| Age, years | 18–39 | 3,840 | 2.9 | 3,770 | 1.3 |
|  | 40–49 | 3,200 | 3.4 | 3,130 | 1.3 |
|  | 50–59 | 4,360 | 3.7 | 4,250 | 1.2 |
|  | 60–69 | 6,070 | 3.5 | 5,940 | 1.3 |
|  | 70+ | 6,340 | 2.7 | 6,230 | 1.0 |
| Race and ethnicity | Hispanic or Latino | 3,730 | 2.9 | 3,660 | 1.1 |
|  | NH Asian | 630 | – | 620 | – |
|  | NH Black | 2,910 | 3.1 | 2,850 | 1.1 |
|  | NH White | 14,580 | 3.2 | 14,290 | 1.2 |
|  | NH other or multiple | 1,280 | 4.7 | 1,240 | – |
|  | Unknown | 680 | 4.4 | 660 | – |

Full cohort: adults with EHR-documented COVID-19 and observation from 365 days before to 455 days after the index. Landmark cohort: those with no Long COVID code by day 90. Counts are rounded up to the next multiple of 10, and percentages use the rounded counts. –: suppressed because a group had 20 or fewer coded patients, or to prevent recovery of a suppressed value from the same table’s totals.

Under the primary survey-comparability assumptions, about 1 in 9 adults who had ever had Long COVID and 1 in 4 who currently had it carried a U09.9 code (Table 2, Figure 3). Women had higher survey prevalence than men but similar coded shares. The women-to-men coding ratio was 0.79 (95% sensitivity interval, 0.65–0.95) for ever having had Long COVID and 0.72 (0.58–0.88) for current Long COVID. The lower fraction for women persisted with within-period survey error and demographic reweighting, and the ever ratio reached 1 only when the relative code-validity-to-prevalence multiplier was about 1.27 (Supplementary Section S2). Hispanic adults had a lower coding fraction than non-Hispanic White adults under the ever definition, and differences across age groups were smaller.

**Figure 3:**
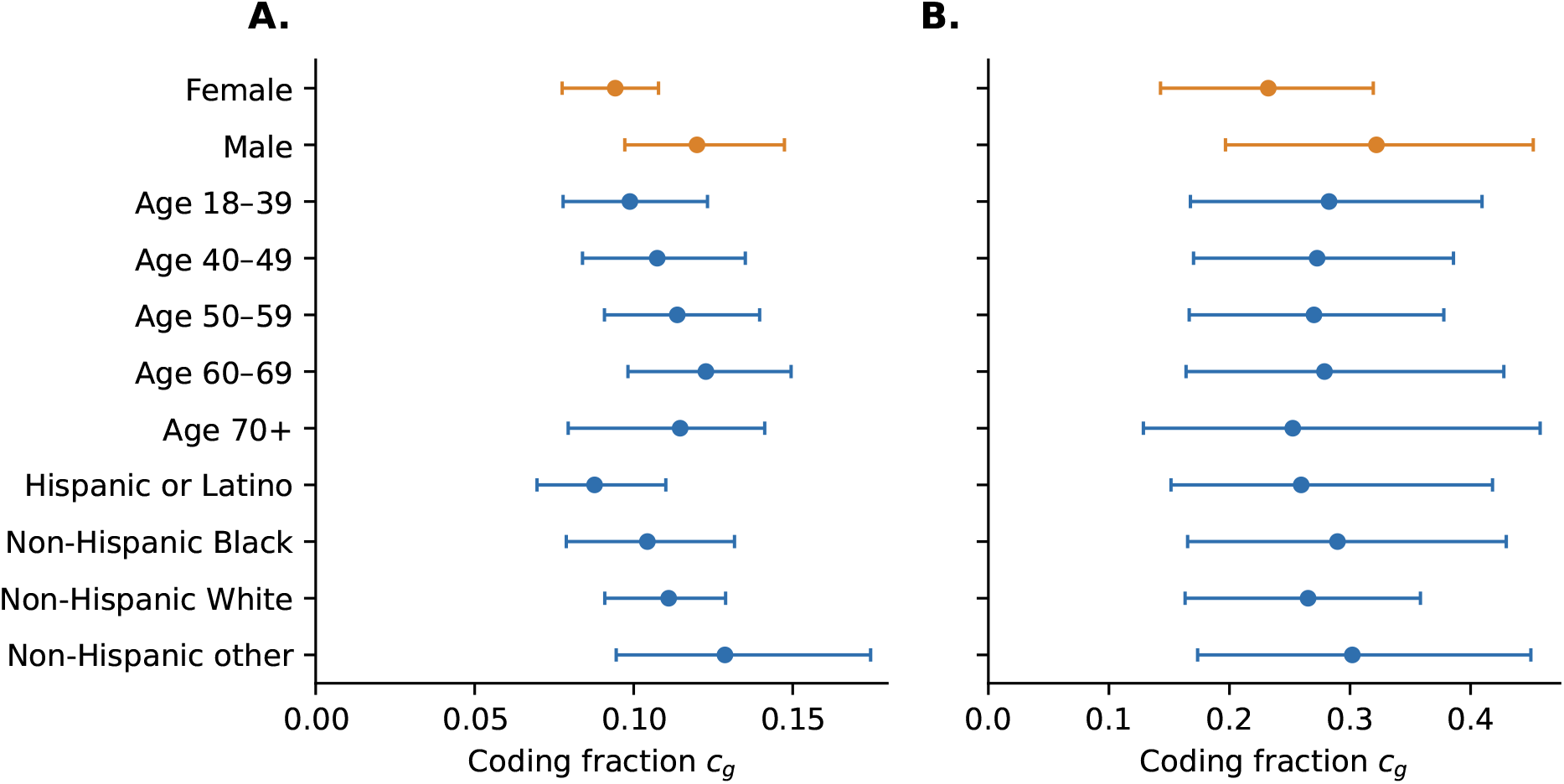
Coding fraction by demographic group. Coded share in days 1–455 divided by survey prevalence, with central 95% sensitivity intervals from survey periods and binomial draws. **A.** Ever having had Long COVID. **B.** Currently having Long COVID. Orange marks the sex groups. Two panels of point estimates with intervals show coding fractions below 1 for every demographic group, under the ever and current survey definitions. Fractions are lower for women than for men under both definitions.

**Table 2:** Coding fraction by demographic group.

|  |  | Coded, % | Ever had Long COVID |  | Currently has Long COVID |  |
| --- | --- | --- | --- | --- | --- | --- |
|  |  |  | Coding fraction | Ratio (95% interval) | Coding fraction | Ratio (95% interval) |
| Sex | Female | 3.2 | 0.094 | 0.79 (0.65–0.95) | 0.232 | 0.72 (0.58–0.88) |
|  | Male | 2.9 | 0.120 | 1 (ref.) | 0.322 | 1 (ref.) |
| Age, years | 18–39 | 2.9 | 0.099 | 0.87 (0.66–1.14) | 0.283 | 1.05 (0.76–1.44) |
|  | 40–49 | 3.4 | 0.107 | 0.94 (0.72–1.22) | 0.273 | 1.01 (0.78–1.32) |
|  | 50–59 | 3.7 | 0.114 | 1 (ref.) | 0.270 | 1 (ref.) |
|  | 60–69 | 3.5 | 0.123 | 1.08 (0.86–1.36) | 0.279 | 1.03 (0.78–1.37) |
|  | 70+ | 2.7 | 0.115 | 1.01 (0.74–1.33) | 0.253 | 0.94 (0.67–1.47) |
| Race and ethnicity | Hispanic or Latino | 2.9 | 0.088 | 0.79 (0.63–0.98) | 0.260 | 0.98 (0.72–1.31) |
|  | NH White | 3.2 | 0.111 | 1 (ref.) | 0.265 | 1 (ref.) |
|  | NH Black | 3.1 | 0.104 | 0.94 (0.73–1.18) | 0.290 | 1.09 (0.82–1.43) |
|  | NH other or multiple | 4.7 | 0.129 | 1.16 (0.84–1.61) | 0.302 | 1.14 (0.78–1.68) |

Coded share in days 1–455 divided by Household Pulse Survey prevalence among adults with prior COVID-19, assuming no false-positive codes and comparable prevalences. Ratios are relative to the indicated reference group. Central 95% sensitivity intervals reflect variation across survey periods and in coded shares.

### 4.2 Prediction from released counts

The patient and site fitting samples retained 40 and 41 of the 102 binary features. Cross-validation within the fitting samples selected NB for the patient partition and LDA with *λ* = 0.6 for the site partition. In the site partition, LDA’s mean cross-validated AUROC exceeded NB’s by 0.005 (0.661 vs 0.657, Supplementary Section S11).

The released models had AUROCs of 0.68 (95% CI, 0.60–0.75) on the patient test and 0.64 (0.55–0.72) at held-out sites (Table 3). At held-out sites, the released model’s AUROC exceeded the binary elastic net’s by 0.049 (95% CI, 0.001–0.104) and equaled the rich elastic net’s point estimate (Figure 4A). On the patient test, it was 0.013 lower than the binary elastic net’s (95% CI, *−*0.070 to 0.049).

**Figure 4:**
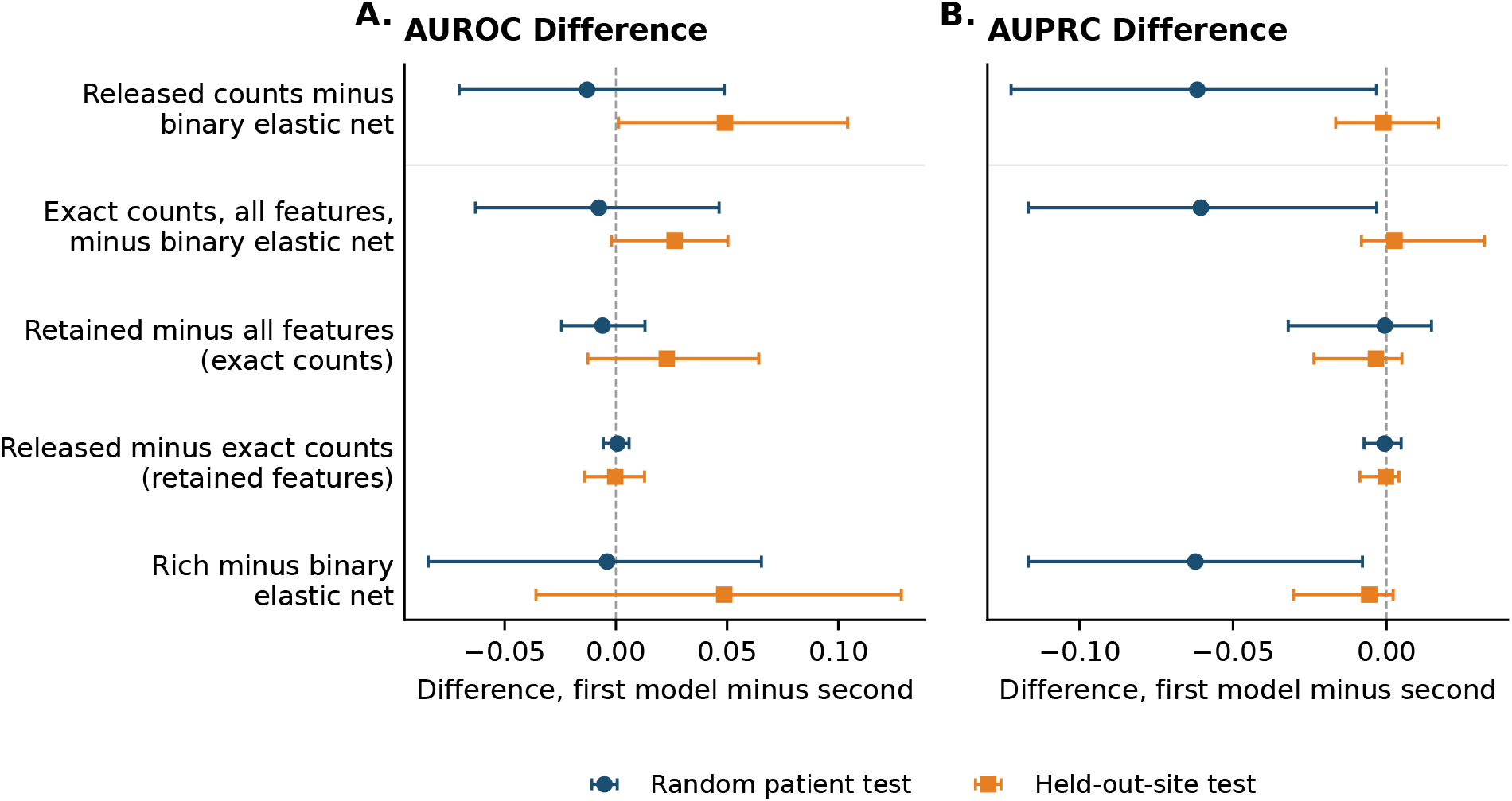
Paired differences between matched models. Each row subtracts the second model from the first, so a negative value means the first, more restricted model performed worse. The top row is the total effect of fitting from released counts instead of participant records. The next three rows divide it into the change of estimator, the exclusion of features that failed the release rule, and count processing. The bottom row compares the two elastic nets. **A.** AUROC. **B.** AUPRC. Points show estimates and bars show 95% CIs from 500 paired, outcome-stratified participant bootstrap replicates. Supplementary Table S15 lists the values. Two forest plots of paired differences, for AUROC and AUPRC, each with five rows and separate markers for the patient test and the held-out-site test. At held-out sites, the released model’s AUROC exceeds the binary elastic net’s, with an interval just above zero. On the patient test, the binary elastic net has the higher AUPRC, and the same gap appears against the exact-count model. Count processing differences sit at zero in both tests.

**Table 3:** Discrimination of matched models for a later U09.9 code.

| Model | Features | AUROC (95% CI) | AUPRC (95% CI) |
| --- | --- | --- | --- |
| *Random patient test* |  |  |  |
| Rich features, elastic net | 123 | 0.688 (0.614, 0.755) | 0.029 (0.019, 0.058) |
| Binary features, elastic net | 102 | 0.692 (0.604, 0.776) | 0.092 (0.027, 0.167) |
| All binary features, exact counts | 102 | 0.684 (0.607, 0.756) | 0.031 (0.018, 0.082) |
| Retained features, exact counts | 40 | 0.678 (0.599, 0.751) | 0.031 (0.019, 0.068) |
| Retained features, released counts | 40 | 0.679 (0.603, 0.753) | 0.030 (0.019, 0.062) |
| *Held-out-site test* |  |  |  |
| Rich features, elastic net | 123 | 0.643 (0.559, 0.718) | 0.017 (0.012, 0.027) |
| Binary features, elastic net | 102 | 0.594 (0.502, 0.682) | 0.022 (0.012, 0.055) |
| All binary features, exact counts | 102 | 0.620 (0.530, 0.701) | 0.025 (0.013, 0.071) |
| Retained features, exact counts | 41 | 0.643 (0.557, 0.717) | 0.022 (0.012, 0.057) |
| Retained features, released counts | 41 | 0.643 (0.553, 0.719) | 0.021 (0.012, 0.055) |

All five models in a partition were fitted to the same patients. Exact- and released-count models use the specification selected by cross-validation within the fitting sample: NB with empirical-Bayes shrinkage in the patient partition and LDA with empirical-Bayes shrinkage and $\lambda = 0.6$ in the site partition. Test sizes: 4,680 patients with 50 later coded (patient test) and 5,760 with 60 (held-out sites), rounded up to the next multiple of 10. AUPRC is average precision. 95% CIs from 500 outcome-stratified participant bootstrap replicates with fixed predictions.

Precision differed on the patient test. The binary elastic net had an AUPRC of 0.09, against 0.03 for the released model (difference 0.062, 95% CI, 0.003–0.122) (Figure 4B). The same gap separated the binary elastic net from the exact-count model with all 102 features (0.060, 95% CI, 0.003–0.117), so it was present before any count was released. The rich elastic net had an AUPRC of 0.03 on the same test, and at held-out sites the released model and the binary elastic net both had AUPRCs of 0.02.

With the estimator and retained features fixed, replacing exact counts with released counts changed AUROC by less than 0.001 in both tests. Excluding the features that failed the release rule changed AUROC by *−*0.006 on the patient test and +0.023 at held-out sites, and both intervals included zero.

### 4.3 Calibration and reproducibility

The released probabilities refer to a later code. At held-out sites, the calibration intercept in the large was *−*0.29 (95% CI, *−*0.58 to *−*0.02), so the predicted odds of a code averaged about 1.3 times the observed odds. On the patient test, the intercept was *−*0.12 (95% CI, *−*0.44 to 0.15). Calibration slopes were 0.89 (95% CI, 0.51–1.19) and 0.83 (0.40–1.18) on the patient and site tests (Supplementary Section S11). Rebuilt from the public archive, the models reproduced the scores and probabilities evaluated in the Workbench to numerical precision. Every retained-feature pair count was released, so neither model needed the independence fallback.

## 5 Discussion

Measured against federal survey prevalence and under the stated comparability assumptions, about 1 in 9 adults who had ever had Long COVID carried its code, and women were coded less completely than men. A model that anyone can rebuild from released counts predicted a later code with AUROCs of 0.68 on a random patient test and 0.64 at held-out sites. At held-out sites, its AUROC exceeded that of an elastic net fitted to participant records of the same patients. On the patient test, that elastic net placed more later-coded patients at the top of its ranking. Count processing itself changed discrimination very little.

The coding fractions agree in direction with the low sensitivity of U09.9 against RECOVER phenotypes and chart review [1, 2]. The lower fraction among women adds a prevalence denominator to earlier reports of demographic differences in Long COVID coding [3, 6–8]. A companion analysis that linked All of Us participants’ self-reported diagnoses to their own records found codes for fewer than half of those reporting ongoing care for chronic fatigue, Lyme disease, or endometriosis [30]. Together with these studies, the survey comparison shows that a low coded prevalence can reflect incomplete documentation more than low disease burden.

Our model forecasts a first code after day 90 from information already in the chart on that day, whereas published All of Us and N3C models use different eligibility rules, feature windows, and reference outcomes [9, 13, 14]. The closest early-prediction study used history and the first 21 days after infection, extended through discharge for longer hospitalizations, and reported cross-site AUROCs of 0.74–0.75 overall [10]. Its AUROC is therefore not a direct benchmark for our cohort. Matching patients, predictors, and outcomes within All of Us instead measures what the release route itself costs.

Releasing the model removed more than half of the binary features, limited the estimator to NB or LDA, and rounded every count, yet none of these restrictions lowered AUROC in both tests, although the intervals are wide. NB and LDA need only class counts, feature-by-outcome tables, and pair counts, so a fixed public release lets anyone rebuild both the coefficients and the probability map, unlike iterative distributed estimation, which needs a continuing connection to the sites [19, 20]. Another institution needs the feature definitions, the reference scorer, and local EHR data mapped to those definitions.

On the patient test, the binary elastic net ranked more later-coded patients at the top than the exact-count model fitted to the same features, so its precision advantage reflects the estimator. The same elastic net with richer features did not show the advantage, and the gap did not appear at held-out sites. With 50 later-coded test patients, a few patients ranked near the top can move AUPRC substantially. We therefore read it as an advantage of this comparator on this test, not as an inherent disadvantage of models fitted from summaries.

If coding errors do not depend on features within disease classes, including a constant false-positive rate, coded-outcome AUROC is a positive affine function of disease AUROC, so models ranked against the code are ranked the same way against disease [27]. Our own data make that global assumption questionable. Coding fractions differed by sex here, and earlier studies found that U09.9 use varies across sites and clinicians [3, 4]. In the companion analysis, the share of participants reporting care for hypertension who had a hypertension code also varied widely across health systems [30]. These differences do not establish that the assumption fails among people with or without Long COVID in the day-90 cohort, because disease status is unobserved and the survey comparison concerns a different population and time window. If coding varies by group, the relation holds at best within groups. We therefore treat it as a conditional interpretation of the model comparison, not as validation of disease ranking. For the same reason, calibration to later coding is distinct from calibration to disease risk. A companion study found that the training label changes which patients a model ranks highly: scores trained on self-reported chronic fatigue or fibromyalgia ranked uncoded self-reporters higher than scores trained on later codes [31].

Because the released model’s discrimination is moderate and it overpredicted at new sites, an institution using it to surface charts for clinician review or research outreach should validate feature extraction locally, recalibrate, and run a silent evaluation before use [32]. The existing SMART on FHIR application shows how a sibling model reaches clinicians [29]. Running the released LDA there would require validating its FHIR inputs against the research definitions.

### 5.1 Limitations

First, the survey comparison depends on code validity and on differences between the cohort and survey populations, and a code anywhere in 455 days may refer to resolved disease. Sensitivity analyses characterize these dependencies, but the coding fractions remain estimates under stated assumptions. Second, the prediction outcome is a recorded code, and the assumptions that link coded-outcome performance to disease are not testable with these data. Third, test events were few, so intervals for AUPRC and calibration are wide. The intervals condition on the fitted models and chosen settings and do not reflect refitting or sampling of new sites. Both test partitions had also served the supplementary analyses of this cohort, so they are not untouched confirmatory samples. Fourth, validation used site groups within one research network, and 12.3% of held-out-site patients also had records from a development site by day 90. Fifth, requiring EHR coverage through day 455 selected patients with sustained observation, and All of Us differs from the US population [33]. Finally, research features and clinical FHIR inputs differ in completeness and representation [34], so the released model needs local feature validation and recalibration before clinical use.

## 6 Conclusion

Compared with federal survey prevalence, Long COVID is substantially and unevenly undercoded in EHRs. A model that anyone can rebuild from counts released by a research enclave predicted later Long COVID documentation with moderate discrimination. Its discrimination was lower than that of a participant-level elastic net on one test and higher on the other, and count processing itself cost little. Matched comparisons make the cost of each release restriction measurable, and local validation of features and calibration is the next step toward chart review.

## Data Availability

All of Us Registered Tier Dataset v9 is available to authorized researchers through the All of Us Researcher Workbench. Household Pulse Survey and Medicare data are public. The aggregate statistics behind every table and figure, the code and a notebook that regenerate them, and the two released models are available at https://github.com/AsiaeeLab/unom-long-covid.

https://www.cdc.gov/nchs/covid19/pulse/long-covid.htm

## Acknowledgments

We gratefully acknowledge All of Us participants for their contributions, without whom this research would not have been possible. We also thank the National Institutes of Health’s All of Us Research Program for making available the participant data examined in this study. The All of Us Research Program is supported by the National Institutes of Health, Office of the Director: Regional Medical Centers: 1 OT2 OD026549; 1 OT2 OD026554; 1 OT2 OD026557; 1 OT2 OD026556; 1 OT2 OD026550; 1 OT2 OD 026552; 1 OT2 OD026553; 1 OT2 OD026548; 1 OT2 OD026551; 1 OT2 OD026555; IAA #: AOD 16037; Federally Qualified Health Centers: HHSN 263201600085U; Data and Research Center: 5 U2C OD023196; Biobank: 1 U24 OD023121; The Participant Center: U24 OD023176; Participant Technology Systems Center: 1 U24 OD023163; Communications and Engagement: 3 OT2 OD023205; 3 OT2 OD023206; and Community Partners: 1 OT2 OD025277; 3 OT2 OD025315; 1 OT2 OD025337; 1 OT2 OD025276. In addition, the All of Us Research Program would not be possible without the partnership of its participants.

## Author contributions

Amir Asiaee (Conceptualization, Methodology, Formal analysis, Data curation, Writing – original draft). Kaveh Aryan (Software, Writing – review and editing).

## Supplementary material

Supplementary material is available at *Journal of the American Medical Informatics Association* online.

## Data availability

All of Us Registered Tier Dataset v9 is available to authorized researchers through the Researcher Workbench. Household Pulse Survey and Medicare data are public. The aggregate statistics behind every table and figure are in the code repository.

## Code availability

A public repository contains the aggregate statistics behind every table and figure, with code and a notebook that regenerate them, the two released models with their feature definitions, calibration maps, reference scorer, and synthetic test cases, and the Workbench scripts that produced the aggregate statistics: https://github.com/AsiaeeLab/unom-long-covid. Running the fitting code on the released counts reproduces the coefficients and calibration maps of the released models, and researchers with Registered Tier access can rerun the Workbench scripts on the same dataset.

## References

[1] Castro VM, Gainer V, Wattanasin N, et al. Characterization and validation of EHR computable phenotypes for Long COVID using patient-reported symptoms: insights from the nationwide RECOVER program. J Am Med Inform Assoc, 33(9):1759–1769, 2026. doi: 10.1093/jamia/ocag115.

[2] Hong C, Wen J, Zhang HG, et al. Label efficient phenotyping for Long COVID using electronic health records. NPJ Digit Med, 8:405, 2025. doi: 10.1038/s41746-025-01617-y.

[3] Pfaff ER, Madlock-Brown C, Baratta JM, et al. Coding long COVID: characterizing a new disease through an ICD-10 lens. BMC Med, 21:58, 2023. doi: 10.1186/s12916-023-02737-6.

[4] Hendrix N, Parikh RV, Taskier M, et al. Heterogeneity of diagnosis and documentation of post-COVID conditions in primary care: A machine learning analysis. PLoS One, 20(5): e0324017, 2025. doi: 10.1371/journal.pone.0324017.

[5] Maripuri M, Dey A, Honerlaw J, et al. Characterization of post–COVID-19 definitions and clinical coding practices: Longitudinal study. Online J Public Health Inform, 16:e53445, 2024. doi: 10.2196/53445.

[6] Butzin-Dozier Z, Ji Y, Deshpande S, et al. Black patients underdiagnosed with Long COVID in the US compared with White patients. Health Aff (Millwood), 45(7):775–781, 2026. doi: 10.1377/hlthaff.2025.00179.

[7] Petranu K, Ford T, Henley D, et al. Exploring Long COVID diagnostic equity by sex, race and ethnicity, and insurance type. Prev Med Rep, 65:103459, 2026. doi: 10.1016/j.pmedr.2026.103459.

[8] Rivelli A, Ording J, Sheehan J, et al. Demographic differences in Long COVID diagnosis across levels of care: Implications for clinical practice. J Gen Intern Med, 2026. doi: 10.1007/s11606-026-10389-4.

[9] Pfaff ER, Girvin AT, Bennett TD, et al. Identifying who has long COVID in the USA: a machine learning approach using N3C data. Lancet Digit Health, 4(7):e532–e541, 2022. doi: 10.1016/S2589-7500(22)00048-6.

[10] Antony B, Blau H, Casiraghi E, et al. Predictive models of long COVID. EBioMedicine, 96: 104777, 2023. doi: 10.1016/j.ebiom.2023.104777.

[11] Zang C, Hou Y, Schenck EJ, et al. Identification of risk factors of Long COVID and predictive modeling in the RECOVER EHR cohorts. Commun Med (Lond), 4(1):130, 2024. doi: 10.1038/s43856-024-00549-0.

[12] Crosskey M, McIntee T, Preiss S, et al. Re-engineering a machine learning phenotype to adapt to the changing COVID-19 landscape: a machine learning modelling study from the N3C and RECOVER consortia. Lancet Digit Health, 7(8):100887, 2025. doi: 10.1016/j.landig.2025.100887.

[13] Pfaff ER, Girvin AT, Crosskey M, et al. De-black-boxing health AI: demonstrating reproducible machine learning computable phenotypes using the N3C-RECOVER Long COVID model in the *All of Us* data repository. J Am Med Inform Assoc, 30(7):1305–1312, 2023. doi: 10.1093/jamia/ocad077.

[14] Guardo C, Zhang X, Gangireddy S, et al. Multi-scale data improves performance of machine learning model for long COVID identification. Commun Med (Lond), 6:389, 2026. doi: 10.1038/s43856-026-01621-7.

[15] Elkan C and Noto K. Learning classifiers from only positive and unlabeled data. In Proceedings of the 14th ACM SIGKDD International Conference on Knowledge Discovery and Data Mining, pages 213–220, 2008. doi: 10.1145/1401890.1401920.

[16] Bekker J and Davis J. Learning from positive and unlabeled data: a survey. Mach Learn, 109 (4):719–760, 2020. doi: 10.1007/s10994-020-05877-5.

[17] Ward G, Hastie T, Barry S, et al. Presence-only data and the EM algorithm. Biometrics, 65 (2):554–563, 2009. doi: 10.1111/j.1541-0420.2008.01116.x.

[18] All of Us Research Program. Frequently asked questions. https://support.researchallofus.org/hc/en-us/articles/52861503885460-Frequently-Asked-Questions, 2026. Accessed October 8, 2026.

[19] Wu Y, Jiang X, Kim J, et al. Grid Binary LOgistic REgression (GLORE): building shared models without sharing data. J Am Med Inform Assoc, 19(5):758–764, 2012. doi: 10.1136/amiajnl-2012-000862.

[20] Gaye A, Marcon Y, Isaeva J, et al. DataSHIELD: taking the analysis to the data, not the data to the analysis. Int J Epidemiol, 43(6):1929–1944, 2014. doi: 10.1093/ije/dyu188.

[21] Kundu R, Salvatore M, Patel KK, et al. Privacy-enhancing sequential learning under heterogeneous selection bias in multi-site electronic health records data. J Am Med Inform Assoc, 33 (9):1679–1692, 2026. doi: 10.1093/jamia/ocag083.

[22] Li KM, Reps JM, Nishimura A, et al. Transfer-learning on federated observational healthcare data for prediction models using Bayesian sparse logistic regression with informed priors. J Am Med Inform Assoc, 33(2):409–423, 2026. doi: 10.1093/jamia/ocaf146.

[23] Ford ND, Slaughter D, Edwards D, et al. Long COVID and significant activity limitation among adults, by age — United States, June 1–13, 2022, to June 7–19, 2023. MMWR Morb Mortal Wkly Rep, 72(32):866–870, 2023. doi: 10.15585/mmwr.mm7232a3.

[24] Collins GS, Moons KGM, Dhiman P, et al. TRIPOD+AI statement: updated guidance for reporting clinical prediction models that use regression or machine learning methods. BMJ, 385:e078378, 2024. doi: 10.1136/bmj-2023-078378.

[25] The All of Us Research Program Investigators. The “All of Us” Research Program. N Engl J Med, 381(7):668–676, 2019. doi: 10.1056/NEJMsr1809937.

[26] National Center for Health Statistics. U.S. Census Bureau, Household Pulse Survey, 2022–2024. Long COVID. https://www.cdc.gov/nchs/covid19/pulse/long-covid.htm. Dataset gsea-w83j. Accessed October 17, 2026.

[27] Jain S, White M, and Radivojac P. Recovering true classifier performance in positive-unlabeled learning. Proc AAAI Conf Artif Intell, 31(1), 2017. doi: 10.1609/aaai.v31i1.10937.

[28] Mandel JC, Kreda DA, Mandl KD, et al. SMART on FHIR: a standards-based, interoperable apps platform for electronic health records. J Am Med Inform Assoc, 23(5):899–908, 2016. doi: 10.1093/jamia/ocv189.

[29] Asiaee A and Aryan K. Design and Technical Evaluation of a Standards-Based System That Delivers Enclave-Derived Case-Finding Models to Patients and Clinicians, 2026. Companion manuscript, submitted.

[30] Asiaee A and Aryan K. Electronic Health Record Coding of Self-Reported Diagnoses in the All of Us Research Program, 2026. Companion manuscript, submitted.

[31] Asiaee A and Aryan K. Self-reported diagnoses as training labels for finding uncoded chronic fatigue, fibromyalgia, and endometriosis in the All of Us Research Program, 2026. Companion manuscript, submitted.

[32] Corbin CK, Maclay R, Acharya A, et al. DEPLOYR: a technical framework for deploying custom real-time machine learning models into the electronic medical record. J Am Med Inform Assoc, 30(9):1532–1542, 2023. doi: 10.1093/jamia/ocad114.

[33] Montgomery BW, Elkasabi M, Brannock MD, et al. Assessing the generalizability of prevalence estimates from the *All of Us* Research Program. Am J Epidemiol, 195(7):1962–1973, 2026. doi: 10.1093/aje/kwag010.

[34] Patterson J, Minto E, Beaton M, et al. A comparison of Fast Healthcare Interoperability Resources and Observational Medical Outcomes Partnership electronic health record data within the All of Us Research Program. J Am Med Inform Assoc, 33(9):1593–1604, 2026. doi: 10.1093/jamia/ocag095.

