## Supplementary Material for "Long COVID undercoding and prediction from releasable electronic health record summaries in All of Us"

#### S1. Cohort selection and decision curve

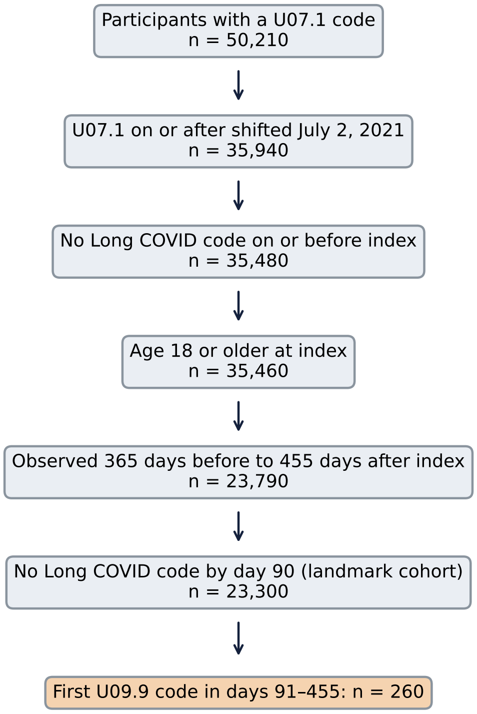


**Figure S1: Cohort selection.** We rounded counts up to a multiple of 10.

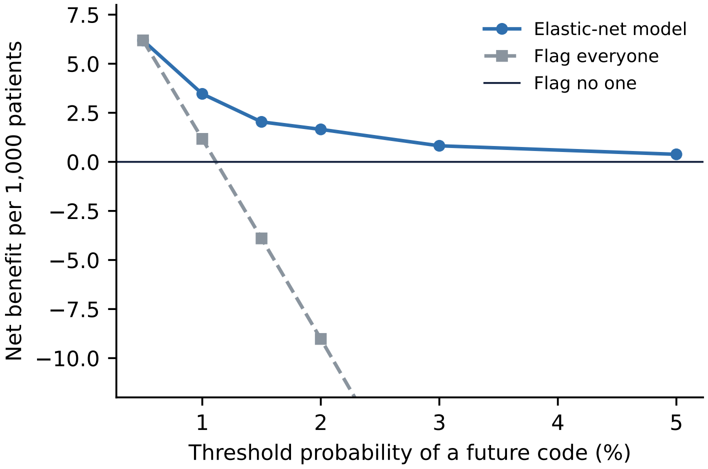


**Figure S2: Decision curve for the coded outcome.** Net benefit of flagging patients whose predicted probability of a subsequent U09.9 code at day 90 exceeds each threshold, compared with flagging everyone or no one, from the out-of-fold elastic-net scores behind Table S10. We computed net benefit from counts rounded up to a multiple of 10. Uncoded patients with Long COVID count as false positives, so the curve describes the coded endpoint, not detection of disease. Above the 2% threshold, the net benefit of flagging everyone falls below the plotted range.

**Table S1: Chart features at the day-90 landmark.** Windows: W0 days −365 to −91; W1 days −90 to −1; W2 days 0 to 30; W3 days 31 to 90. "New" features require a match in W2 or W3 and none in W0 or W1. ICD-10-CM codes match by prefix, without dots. Encounter classes: IMP inpatient, EMER emergency, AMB all other visits except pharmacy and laboratory visits. The public research package defines all 102 inputs and the 40 and 41 fitted features in the patient and source instances, respectively. Medication features use ingredient descendants in RxNorm and RxNorm Extension; the supplied public RxNorm code list is a subset of that resolver.

| Feature | Group | Definition | Window |
|---|---|---|---|
| Age 18-39 | Demographics | Age at index 18-39 years | at index |
| Age 40-49 | Demographics | Age at index 40-49 years | at index |
| Age 50-59 | Demographics | Age at index 50-59 years | at index |
| Age 60-69 | Demographics | Age at index 60-69 years | at index |
| Age 70+ | Demographics | Age at index 70+ years | at index |
| Female | Demographics | Female sex at birth | at index |
| Hospitalized around the COVID diagnosis | Acute illness | Any encounter of class IMP | days −3 to 30 |
| Emergency visit in the first month | Acute illness | Any encounter of class EMER | days 0 to 30 |
| COVID pneumonia | Acute illness | ICD-10-CM J1282 | days 0 to 30 |
| Ventilation or intubation | Acute illness | CPT 94002, 94003, 31500; ICD-10-PCS 5A1935Z, 5A1945Z, 5A1955Z | days 0 to 30 |
| 4+ outpatient visits in days −365 to −91 | Health care use | 4 or more encounters of class AMB | W0 |
| 2+ outpatient visits in days 31-90 | Health care use | 2 or more encounters of class AMB | W3 |
| 4+ outpatient visits in days 31-90 | Health care use | 4 or more encounters of class AMB | W3 |
| Emergency visit in days 31-90 | Health care use | Any encounter of class EMER | W3 |
| Hospital stay in days 31-90 | Health care use | Any encounter of class IMP | W3 |
| Visit rate after COVID at least double the baseline rate | Health care use | Encounters of class AMB or EMER per day in W3 at least 2 times the rate in W0, or any in W3 with none in W0 | W3 vs W0 |
| New: fatigue or malaise | New symptoms | ICD-10-CM R53, G933 | W2+W3 (none in W0+W1) |
| New: shortness of breath | New symptoms | ICD-10-CM R060 | W2+W3 (none in W0+W1) |
| New: cough | New symptoms | ICD-10-CM R05 | W2+W3 (none in W0+W1) |
| New: chest pain | New symptoms | ICD-10-CM R07 | W2+W3 (none in W0+W1) |
| New: palpitations, tachycardia, POTS, or orthostatic hypotension | New symptoms | ICD-10-CM R000, R002, I498, G90A, I951 | W2+W3 (none in W0+W1) |
| New: cognitive symptoms (brain fog, memory) | New symptoms | ICD-10-CM R41, G3184 | W2+W3 (none in W0+W1) |
| New: headache | New symptoms | ICD-10-CM R51, G44 | W2+W3 (none in W0+W1) |
| New: dizziness or fainting | New symptoms | ICD-10-CM R42, R55 | W2+W3 (none in W0+W1) |

| Feature | Group | Definition | Window |
| --- | --- | --- | --- |
| New: loss or change of smell or taste | New symptoms | ICD-10-CM R43 | W2+W3 (none in W0+W1) |
| New: sleep disturbance | New symptoms | ICD-10-CM G470, G471, G478, G479, F51 | W2+W3 (none in W0+W1) |
| New: anxiety or depression | New symptoms | ICD-10-CM F41, F32, F33, F43 | W2+W3 (none in W0+W1) |
| New: muscle or joint pain | New symptoms | ICD-10-CM M791, M255, M796, M797 | W2+W3 (none in W0+W1) |
| New: gastrointestinal symptoms | New symptoms | ICD-10-CM R10, R11, R14, R19, K58, K59 | W2+W3 (none in W0+W1) |
| New: tingling, numbness, or neuropathy | New symptoms | ICD-10-CM R20, G629 | W2+W3 (none in W0+W1) |
| New: hair loss | New symptoms | ICD-10-CM L659 | W2+W3 (none in W0+W1) |
| New: fever | New symptoms | ICD-10-CM R50 | W2+W3 (none in W0+W1) |
| Two or more new symptom types | New symptoms | 2 or more of the 16 new-symptom features above | W2+W3 (none in W0+W1) |
| Four or more new symptom types | New symptoms | 4 or more of the 16 new-symptom features above | W2+W3 (none in W0+W1) |
| History: asthma | Prior conditions | ICD-10-CM J45 | W0+W1 |
| History: COPD | Prior conditions | ICD-10-CM J44 | W0+W1 |
| History: diabetes | Prior conditions | ICD-10-CM E10, E11 | W0+W1 |
| History: obesity | Prior conditions | ICD-10-CM E66, Z683, Z684 | W0+W1 |
| History: hypertension | Prior conditions | ICD-10-CM I10 | W0+W1 |
| History: depression | Prior conditions | ICD-10-CM F32, F33 | W0+W1 |
| History: anxiety | Prior conditions | ICD-10-CM F41 | W0+W1 |
| History: migraine | Prior conditions | ICD-10-CM G43 | W0+W1 |
| History: fibromyalgia | Prior conditions | ICD-10-CM M797 | W0+W1 |
| History: ME/CFS | Prior conditions | ICD-10-CM G933 | W0+W1 |
| History: chronic pain | Prior conditions | ICD-10-CM G89 | W0+W1 |
| History: sleep apnea | Prior conditions | ICD-10-CM G4733 | W0+W1 |
| History: autoimmune | Prior conditions | ICD-10-CM M05, M06, M32, L40, K50, K51, M45 | W0+W1 |
| History: hypothyroid | Prior conditions | ICD-10-CM E03 | W0+W1 |
| History: heart disease | Prior conditions | ICD-10-CM I25, I50 | W0+W1 |
| Echocardiogram | Tests after COVID | CPT 93303-93308, 93312-93318, 93350-93352 | W2+W3 |

| Feature | Group | Definition | Window |
|---|---|---|---|
| Holter or event monitor | Tests after COVID | CPT 93224-93229, 93241-93248, 93268-93272 | W2+W3 |
| Pulmonary function test | Tests after COVID | CPT 94010, 94060, 94070, 94726-94729 | W2+W3 |
| Chest CT | Tests after COVID | CPT 71250, 71260, 71270, 71271, 71275 | W2+W3 |
| Chest X-ray | Tests after COVID | CPT 71045-71048 | W2+W3 |
| Brain MRI | Tests after COVID | CPT 70551-70553 | W2+W3 |
| Tilt-table test | Tests after COVID | CPT 93660 | W2+W3 |
| Sleep study | Tests after COVID | CPT/HCPCS 95806-95811, G0398, G0399, G0400 | W2+W3 |
| Six-minute walk test | Tests after COVID | CPT 94618 | W2+W3 |
| D-dimer test | Tests after COVID | LOINC measurement concepts whose name contains "D-dimer" (12 codes) | W2+W3 |
| Troponin test | Tests after COVID | LOINC measurement concepts whose name contains "Troponin" (25 codes) | W2+W3 |
| BNP or NT-proBNP test | Tests after COVID | LOINC measurement concepts whose name contains "Natriuretic peptide" (10 codes) | W2+W3 |
| Thyroid (TSH) test | Tests after COVID | LOINC measurement concepts whose name contains "Thyrotropin" (98 codes) | W2+W3 |
| Ferritin test | Tests after COVID | LOINC measurement concepts whose name contains "Ferritin" (9 codes) | W2+W3 |
| C-reactive protein test | Tests after COVID | LOINC measurement concepts whose name contains "C reactive protein" (13 codes) | W2+W3 |
| Sedimentation rate (ESR) test | Tests after COVID | LOINC measurement concepts whose name contains "Erythrocyte sedimentation rate" (7 codes) | W2+W3 |
| Vitamin D test | Tests after COVID | LOINC measurement concepts whose name contains "25-hydroxyvitamin D" (22 codes) | W2+W3 |
| Vitamin B12 test | Tests after COVID | LOINC measurement concepts whose name contains "Cobalamin" (38 codes) | W2+W3 |
| Antinuclear antibody (ANA) test | Tests after COVID | LOINC measurement concepts whose name contains "Nuclear Ab" (277 codes) | W2+W3 |
| New beta blocker | New medications | RxNorm ingredients propranolol, metoprolol, atenolol, bisoprolol, nadolol, carvedilol with their descendant drugs in RxNorm or RxNorm Extension | W2+W3 (none in W0+W1) |

| Feature | Group | Definition | Window |
|---|---|---|---|
| New POTS medication | New medications | RxNorm ingredients ivabradine, midodrine, fludrocortisone with their descendant drugs in RxNorm or RxNorm Extension | W2+W3 (none in W0+W1) |
| New SSRI or SNRI antidepressant | New medications | RxNorm ingredients sertraline, fluoxetine, citalopram, escitalopram, paroxetine, venlafaxine, duloxetine, desvenlafaxine with their descendant drugs in RxNorm or RxNorm Extension | W2+W3 (none in W0+W1) |
| New gabapentin or pregabalin | New medications | RxNorm ingredients gabapentin, pregabalin with their descendant drugs in RxNorm or RxNorm Extension | W2+W3 (none in W0+W1) |
| New inhaler | New medications | RxNorm ingredients albuterol, fluticasone, budesonide, mometasone, beclomethasone, tiotropium, formoterol, salmeterol with their descendant drugs in RxNorm or RxNorm Extension | W2+W3 (none in W0+W1) |
| New oral or injected steroid | New medications | RxNorm ingredients prednisone, methylprednisolone, dexamethasone, prednisolone with their descendant drugs in RxNorm or RxNorm Extension | W2+W3 (none in W0+W1) |
| New antihistamine or famotidine | New medications | RxNorm ingredients cetirizine, loratadine, fexofenadine, levocetirizine, famotidine with their descendant drugs in RxNorm or RxNorm Extension | W2+W3 (none in W0+W1) |
| New montelukast | New medications | RxNorm ingredient montelukast with its descendant drugs in RxNorm or RxNorm Extension | W2+W3 (none in W0+W1) |
| New naltrexone | New medications | RxNorm ingredient naltrexone with its descendant drugs in RxNorm or RxNorm Extension | W2+W3 (none in W0+W1) |
| New stimulant or wakefulness drug | New medications | RxNorm ingredients modafinil, armodafinil, methylphenidate, amphetamine, dextroamphetamine, lisdexamfetamine with their descendant drugs in RxNorm or RxNorm Extension | W2+W3 (none in W0+W1) |
| New sleep medication | New medications | RxNorm ingredients zolpidem, trazodone, eszopiclone with their descendant drugs in RxNorm or RxNorm Extension | W2+W3 (none in W0+W1) |
| New opioid | New medications | RxNorm ingredients oxycodone, hydrocodone, tramadol, morphine with their descendant drugs in RxNorm or RxNorm Extension | W2+W3 (none in W0+W1) |
| Fatigue or malaise (coded by day 90) | Symptoms after COVID | ICD-10-CM R53, G933 | days 0 to 90 |

| Feature | Group | Definition | Window |
|---|---|---|---|
| Shortness of breath (coded by day 90) | Symptoms after COVID | ICD-10-CM R060 | days 0 to 90 |
| Cough (coded by day 90) | Symptoms after COVID | ICD-10-CM R05 | days 0 to 90 |
| Chest pain (coded by day 90) | Symptoms after COVID | ICD-10-CM R07 | days 0 to 90 |
| Palpitations, tachycardia, POTS, or orthostatic hypotension (coded by day 90) | Symptoms after COVID | ICD-10-CM R000, R002, I498, G90A, I951 | days 0 to 90 |
| Cognitive symptoms (brain fog, memory) (coded by day 90) | Symptoms after COVID | ICD-10-CM R41, G3184 | days 0 to 90 |
| Headache (coded by day 90) | Symptoms after COVID | ICD-10-CM R51, G44 | days 0 to 90 |
| Dizziness or fainting (coded by day 90) | Symptoms after COVID | ICD-10-CM R42, R55 | days 0 to 90 |
| Loss or change of smell or taste (coded by day 90) | Symptoms after COVID | ICD-10-CM R43 | days 0 to 90 |
| Sleep disturbance (coded by day 90) | Symptoms after COVID | ICD-10-CM G470, G471, G478, G479, F51 | days 0 to 90 |
| Anxiety or depression (coded by day 90) | Symptoms after COVID | ICD-10-CM F41, F32, F33, F43 | days 0 to 90 |
| Muscle or joint pain (coded by day 90) | Symptoms after COVID | ICD-10-CM M791, M255, M796, M797 | days 0 to 90 |
| Gastrointestinal symptoms (coded by day 90) | Symptoms after COVID | ICD-10-CM R10, R11, R14, R19, K58, K59 | days 0 to 90 |
| Tingling, numbness, or neuropathy (coded by day 90) | Symptoms after COVID | ICD-10-CM R20, G629 | days 0 to 90 |
| Hair loss (coded by day 90) | Symptoms after COVID | ICD-10-CM L659 | days 0 to 90 |
| Fever (coded by day 90) | Symptoms after COVID | ICD-10-CM R50 | days 0 to 90 |
| 2+ symptom types coded in days 31-90 | Symptoms after COVID | 2 or more of the 16 symptom clusters coded (new or not) | days 31 to 90 |
| 3+ symptom types coded in days 31-90 | Symptoms after COVID | 3 or more of the 16 symptom clusters coded (new or not) | days 31 to 90 |
| COVID-19 still coded at a follow-up visit (days 31-90) | Coding after COVID | ICD-10-CM U071 | days 31 to 90 |
| Sequelae of infection coded after COVID | Coding after COVID | ICD-10-CM B948 | days 0 to 90 |
| Personal history of COVID-19 coded (days 31-90) | Coding after COVID | ICD-10-CM Z8616 | days 31 to 90 |
| Visit in days 61-90 | Health care use | Any visit except types containing inpatient, pharmacy, or laboratory | days 61 to 90 |

#### S2. Sensitivity of the coding fraction

The primary coding fraction assumes no false-positive codes ($v_g = 1$) and equal prevalence in the All of Us cohort and the survey population ($\kappa_g = 1$). Under Equation 1 of the main text, the women-to-men coding ratio equals the primary ratio multiplied by $(v_F/v_M)/(\kappa_F/\kappa_M)$. Figure S3 shows the ratio over a grid of both quantities. The point estimate reaches 1 only when $(v_F/v_M)/(\kappa_F/\kappa_M)$ is at least 1.27 (ever definition) or 1.39 (current definition). The upper interval bound reaches 1 at 1.05 or 1.13. When we added each survey period's sampling error to the bootstrap, the interval for women widened from 0.65–0.95 to 0.65–0.96 (ever) and from 0.58–0.88 to 0.56–0.92 (current). For composition reweighting, we used nine HPS public-use waves (weeks 46, 48, 50, 51, 54, 56, 58, 60, and 62). Their unweighted-composition prevalence by sex reproduced the published series for the same weeks to within 0.1 percentage point. After we raked within each sex to the All of Us cohort's age-band and race-and-ethnicity margins (computed from counts rounded up to a multiple of 10), mean prevalence changed by factors of 0.98 (women) and 0.97 (men) for ever having Long COVID and 1.03 and 1.09 for current Long COVID. This adjusts composition only. Differences in health and health-care use between volunteers and survey respondents, and between EHR-documented and self-reported COVID-19, remain in $\kappa_g$. To gauge the size of a sex difference in $\kappa$, we estimated it for two conditions that are coded reliably and measured in a national survey. Among adults with at least one EHR visit in each of two consecutive years who completed the All of Us Personal and Family Health History survey, the share with any ICD-10-CM code among participants reporting a clinician's diagnosis was 0.78 (women) and 0.82 (men) for hypertension and 0.81 and 0.88 for type 2 diabetes. We standardized each sex's coded share to the age distribution of US adults (NHANES 2021–2023 interview weights) and divided it by that share to estimate the cohort's diagnosed prevalence. Dividing this prevalence by the prevalence of an ever-told diagnosis in the CDC/NCHS 2023 National Health Interview Survey (Sample Adult public-use file, survey weights) gives the cohort-to-national prevalence ratio. The ratios were 1.43 (women) and 1.49 (men) for hypertension and 2.42 and 2.64 for type 2 diabetes, so $\kappa_F/\kappa_M$ was 0.96 and 0.92 (0.94 and 0.89 when also standardized to race and ethnicity). These conditions are not Long COVID, and $\kappa$ differed by condition, so the values indicate the plausible size of a sex difference rather than its value for Long COVID. With $v_F = v_M$, the women-to-men coding ratio reaches 1 only if $\kappa_F/\kappa_M$ is 0.79 or lower (ever) or 0.72 or lower (current). When we changed the weights that combine HPS age groups (0.3 to 0.7 for ages 18–29 within 18–39, and 0.5 to 0.8 for 70–79 within 70 or older), age-band coding ratios changed by 0.03 or less.

*Index dates before U09.9 was available.* For index dates from July 2 to September 29, 2021 (shifted), some of days 1 to 90 can fall before U09.9 was introduced on October 1, 2021, which could lower the coded share in the coding-fraction cohort. When we restricted the cohort to shifted index dates on or after September 30, 2021, it had 18,830 adults, and their coded share in days 1 to 455 was 2.8%. The restriction lowered the coded share among women, in every age band, and in every racial and ethnic group shown except non-Hispanic Black adults, whose share was similar: adults with earlier index dates were coded more often, not less, so including them did not lower the estimated coding fractions.

#### S3. Related post-infectious conditions

ME/CFS and postviral fatigue (G93.3x), POTS and dysautonomia (G90.A, G90.9), fibromyalgia (M79.7), mast cell disorders (D89.4x), and chronic fatigue (R53.82) share symptoms and coding barriers with Long COVID. At day 90, incident codes in days 91 to 455 numbered 80 for ME/CFS

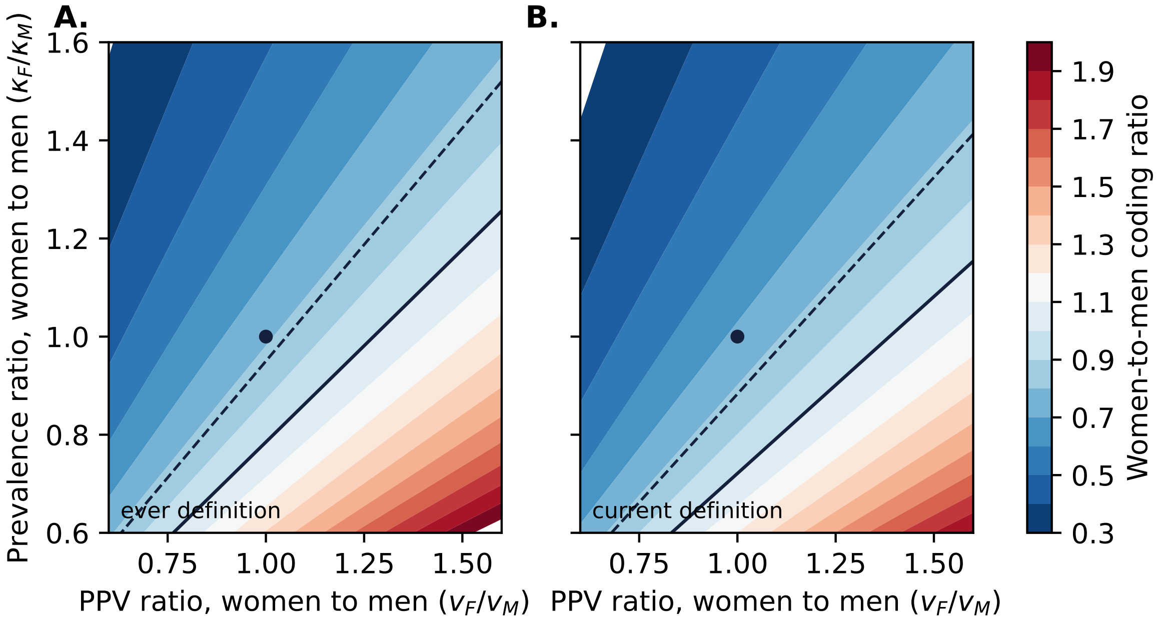


**Figure S3: Women-to-men coding ratio under differences in code validity and survey comparability.** Shading shows the ratio as the women-to-men ratio of positive predictive values ($v_F/v_M$) and of cohort-to-survey prevalence ($\kappa_F/\kappa_M$) vary. The solid line marks where the point estimate equals 1, the dashed line where the upper 95% bound equals 1, and the dot the primary analysis. **A.** Ever-had definition. **B.** Current definition.

or postviral fatigue (30 of them also coded with Long COVID), 80 for POTS or dysautonomia, 210 for fibromyalgia, 240 for chronic fatigue, and 20 or fewer for mast cell disorders, which we excluded. We compared a single-task logistic model for Long COVID with a multi-task logistic model in which each condition $k$ has weights $\beta_k = \beta_0 + \delta_k$, with the deviations $\delta_k$ penalized toward the shared weights, and with a model of a pooled outcome (Long COVID or any related code). We fit the standardization inside each training fold, and each condition had its own intercept. With identical folds, AUROC for Long COVID was 0.662 (0.632–0.697) for the single-task model, 0.672 (0.643–0.702) for the multi-task model, and 0.666 (0.632–0.699) for the pooled outcome. Borrowing strength from related conditions changed discrimination little.

#### S4. Subgroups and analysis provenance

**Table S2: Discrimination, calibration, and flagging of the day-90 elastic net by group.** Out-of-fold predictions from five-fold cross-validation with folds different from those of Table S10, so the overall AUROC differs slightly from the Table S10 estimate of 0.680. We rounded counts up to a multiple of 10 and computed the observed rate from the rounded counts, so observed-to-predicted ratios are approximate. Flagged: share of the group among the top 5% of all scores. We do not show groups with 20 or fewer coded patients, whose estimates are unstable. –: withheld for men, because with the overall row these values would let a reader recover the suppressed values for other or unknown sex (complementary suppression).

| | Group | Patients | Coded | AUROC (95% CI) | Observed/predicted | Flagged at 5% workload, % |
|---|---|---|---|---|---|---|
| All patients | | 23,300 | 260 | 0.688 (0.650–0.724) | 1.01 | 5.0 |
| Sex | Female | 14,550 | 170 | 0.697 (0.657–0.739) | 1.03 | 5.4 |
| | Male | 8,560 | – | 0.667 (0.603–0.719) | – | – |
| Race and ethnicity | Hispanic or Latino | 3,660 | 40 | 0.676 (0.590–0.747) | 1.02 | 4.4 |
| | NH Black | 2,850 | 30 | 0.697 (0.569–0.808) | 0.85 | 7.0 |
| | NH White | 14,290 | 170 | 0.700 (0.658–0.739) | 1.11 | 4.8 |
| Age, years | 18–39 | 3,770 | 50 | 0.759 (0.675–0.837) | 1.27 | 4.5 |
| | 40–49 | 3,130 | 40 | 0.541 (0.446–0.622) | 1.14 | 5.1 |
| | 50–59 | 4,250 | 50 | 0.657 (0.576–0.733) | 1.02 | 5.6 |
| | 60–69 | 5,940 | 80 | 0.733 (0.674–0.781) | 1.16 | 5.7 |
| | 70+ | 6,230 | 60 | 0.685 (0.616–0.756) | 0.94 | 4.5 |

**Table S3: Exploratory score adjustment.** We divided the day-90 elastic-net out-of-fold scores of women by the women-to-men coding ratio $r$ (men and other sex unchanged) and computed shares from counts rounded up to a multiple of 10.

| Ranking | Definition | Ratio $r$ | Women in top 5%, % | Recall of coded in top 5%, % | AUROC |
|---|---|---|---|---|---|
| Unadjusted | – | 1.00 | 67 | 19 | 0.688 |
| Adjusted, point ratio | ever | 0.79 | 75 | 23 | 0.684 |
| Adjusted, lower bound | ever | 0.65 | 80 | 19 | 0.678 |
| Adjusted, upper bound | ever | 0.95 | 69 | 19 | 0.688 |
| Adjusted, point ratio | current | 0.72 | 78 | 19 | 0.681 |
| Adjusted, lower bound | current | 0.58 | 84 | 19 | 0.673 |
| Adjusted, upper bound | current | 0.88 | 72 | 19 | 0.687 |
| Cohort | – | – | 62 | – | – |

**Table S4: Cohort, features, evaluation, and model behind each result.** "Matrix A" is the day-90 matrix of 102 features that we built once for the additional prediction analysis, with ICD-10-CM codes from the condition and observation tables. "Rebuilt" matrices use the same code sources, and we recomputed them at each landmark from the same registry with windows ending at the landmark (we defined the recent-symptom and late-visit windows relative to the landmark), so day-90 estimates differ slightly between the two. B: bootstrap replicates.

| Result | Cohort and outcome | Features | Evaluation | Model |
|---|---|---|---|---|
| Coding fractions (main-text Table 2, Figure 3, Section S2) | 23,790 adults, U09.9 in days 1–455 | none | survey periods and binomial draws, B = 5,000 | none |
| Additional day-90 prediction and documentation associations (Table S10, Figure S4) | 23,300 uncoded at day 90, first U09.9 in days 91–455 | matrix A | 5-fold stratified CV (B = 200), 18 held-out sites pooled, and documentation ratios (B = 200) | M0–M4 |
| Landmark tasks (Figure S5) | rebuilt at each landmark (early landmarks restricted by index date), first U09.9 in days $L + 1$–455 | rebuilt | 5-fold stratified CV, B = 200 | elastic net |
| Day-30 vs day-180 scores (Section S10) | eligible at day 180 and in the day-30 cohort, first U09.9 in days 181–455 | rebuilt | out-of-fold scores of two separately trained models, B = 200 | elastic net |
| Subgroups (Table S2) | as day-90 prediction | matrix A | 5-fold stratified CV (folds different from Table S10), B = 200 | elastic net |
| Score adjustment, workload comparison (Table S3, Section S10) | as day-90 prediction | matrix A | 5-fold stratified CV (folds of Table S2) | elastic net, M1, symptom-count rule |
| Decision curve (Figure S2) | as day-90 prediction | matrix A | out-of-fold scores of Table S10 | elastic net |
| Related conditions (Section S3) | landmark 90, incident related codes | rebuilt | 5-fold stratified CV (identical folds), B = 200 | logistic, multi-task |
| Application model (Table S11) | as day-90 prediction, split at random into 80% training and 20% validation (seed 20260929) | matrix A, 57 eligible features | validation split, B = 200 | naive Bayes, linear discriminants |
| Separate 41-feature instance (Section S10) | 18 held-out sites, with training sites split 70/30 into fit and calibration | matrix A, 41 eligible features | held-out sites, paired B = 500 | naive Bayes (shrinkage), elastic net |
| Utilization before documentation (Section S10) | later-coded patients vs uncoded in matched strata | visit and test counts | bootstrap | none |

| Result | Cohort and outcome | Features | Evaluation | Model |
| --- | --- | --- | --- | --- |
| Binary vs richer features (Section S8) | 23,300 uncoded at day 90, first U09.9 in days 91–455 | matrix A and rich counts | 5-fold stratified CV and 18 held-out sites, B = 200 | elastic nets, gradient boosting |
| Index-date restriction (Section S2) | 18,830 adults with shifted index on or after September 30, 2021, U09.9 in days 1–455 | none | none | none |
| Laboratory results and vital signs (Section S6) | as day-90 prediction | matrix A plus laboratory-result and vital-sign features | 5-fold stratified CV (folds of Table S2) and 18 held-out sites, B = 200 | elastic net |
| Alternative model families (Section S7, Table S6) | as day-90 prediction | matrix A | nested 5-fold CV and 18 held-out sites, B = 200 | boosted trees, random forest, decision tree, elastic net |
| Development and evaluation populations, source grouping (Tables S8 and S9) | as day-90 prediction, both splits | selected features of matrix A | none | none |

#### S5. Summary-count fitting and interpretation of coded outcomes

**Population and targets.** All probabilities below refer to the day-90 eligible population: adults with no Long COVID code by that landmark. Let $X$ contain features available by day 90, $C$ indicate a first recorded U09.9 code in days 91–455, and $Y$ denote the corresponding latent Long COVID status during follow-up. Write $q(x) = P(C = 1 \mid X = x)$, $\eta(x) = P(Y = 1 \mid X = x)$, and $r(x) = \beta_0 + w^\mathsf{T} x$ for a fitted linear score. The Household Pulse Survey comparison concerns full-cohort, group-level prevalence and coding over a different window. It identifies neither $P(Y = 1)$ nor individual coding probabilities in this landmark population.

**Exact summaries and linear scores.** For summary-count fitting, restrict to $X \in \{0, 1\}^d$. For a fitting sample $\{(x_i, c_i)\}_{i=1}^n$, define

$$n_c = \sum_i \mathbf{1}\{c_i = c\}, \qquad t_{cj} = \sum_{i:c_i=c} x_{ij}, \qquad Q_{jk} = \sum_i x_{ij} x_{ik}, \qquad c \in \{0, 1\}.$$

Here $n = n_0 + n_1$, $\widehat{\pi} = n_1/n$, $\widehat{p}_{cj} = t_{cj}/n_c$, $\mu_c = (\widehat{p}_{c1}, \ldots, \widehat{p}_{cd})^\mathsf{T}$, and $m = (n_0\mu_0 + n_1\mu_1)/n$. The four cells for feature $j$ are $t_{1j}, n_1 - t_{1j}, t_{0j}, n_0 - t_{0j}$. Since features are binary, $Q_{jj} = t_{0j} + t_{1j}$. Under conditional feature independence given $C$, Bernoulli naive Bayes (NB) has fitted log odds

$$\log \frac{\widehat{P}(C = 1 \mid x)}{\widehat{P}(C = 0 \mid x)} = \log \frac{n_1}{n_0} + \sum_j \left[ x_j \log \frac{\widehat{p}_{1j}}{\widehat{p}_{0j}} + (1 - x_j) \log \frac{1 - \widehat{p}_{1j}}{1 - \widehat{p}_{0j}} \right].$$

Collecting terms gives

$$w_j = \log \frac{\widehat{p}_{1j}(1 - \widehat{p}_{0j})}{\widehat{p}_{0j}(1 - \widehat{p}_{1j})}, \qquad \beta_0 = \log \frac{n_1}{n_0} + \sum_j \log \frac{1 - \widehat{p}_{1j}}{1 - \widehat{p}_{0j}}.$$

These expressions assume both classes and all four feature cells are positive. Our export procedure keeps features only when all four cells exceed 20. That screening choice, rather than a restriction inherent to binary features, avoids zero and rare cells. Other specifications could use smoothing.

Empirical-Bayes shrinkage is an additional approximation, separate from NB. Using the normal working model $w_j \mid \theta_j \sim N(\theta_j, v_j)$, $\theta_j \sim N(0, \tau^2)$, the implementation sets

$$v_j = \frac{1}{t_{1j}} + \frac{1}{n_1 - t_{1j}} + \frac{1}{t_{0j}} + \frac{1}{n_0 - t_{0j}}, \qquad \widehat{\tau}^2 = \max\left\{0, \frac{1}{d} \sum_j (w_j^2 - v_j)\right\}, \qquad \widetilde{w}_j = \frac{\widehat{\tau}^2}{\widehat{\tau}^2 + v_j} w_j.$$

It uses the centered intercept $\widetilde{\beta}_0 = \log(n_1/n_0) - \widetilde{w}^\mathsf{T} m$, which centers the score at the sample feature mean but does not itself calibrate probabilities. It is not the unmodified NB intercept.

For linear discriminant analysis (LDA), set $\Delta = \mu_1 - \mu_0$. Expanding the within-class scatter gives

$$\begin{aligned} \widehat{\Sigma}_W &= \frac{1}{n} \sum_{c=0}^{1} \sum_{i:c_i=c} (x_i - \mu_c)(x_i - \mu_c)^\mathsf{T} \\ &= \frac{Q}{n} - \sum_{c=0}^{1} \frac{n_c}{n} \mu_c \mu_c^\mathsf{T} = \frac{Q}{n} - m m^\mathsf{T} - \widehat{\pi}(1 - \widehat{\pi}) \Delta \Delta^\mathsf{T}. \end{aligned}$$

The divisor is $n$, the maximum-likelihood convention, rather than $n-2$. Let $D$ be the diagonal matrix with the diagonal of $\widehat{\Sigma}_W$. For $0 \le \lambda \le 1$, diagonal shrinkage gives

$$\Sigma_\lambda = (1-\lambda)\widehat{\Sigma}_W + \lambda D, \qquad w_\lambda = \Sigma_\lambda^{-1}\Delta, \qquad \beta_{0\lambda} = \log\frac{n_1}{n_0} - \tfrac{1}{2}w_\lambda^{\mathsf{T}}(\mu_1+\mu_0),$$

provided the covariance used is positive definite. Gaussian LDA is a working model for these binary features. The additional LDA empirical-Bayes variant applies the same shrinkage rule to $\Delta_j/\sqrt{D_{jj}}$, with variance $[\widehat{p}_{1j}(1-\widehat{p}_{1j})/n_1 + \widehat{p}_{0j}(1-\widehat{p}_{0j})/n_0]/D_{jj}$, then substitutes the resulting unstandardized difference for $\Delta$ in the weight equation, retaining the displayed intercept formula.

Thus exact summaries reproduce matching row-based NB and LDA fits, including the same feature set, denominators, smoothing and shrinkage settings. Protected summaries need not: suppression and rounding can destroy moment consistency. Our implementation subtracts 4.5 from each positive rounded count, a heuristic correction rather than recovery of exact counts, and replaces unavailable pair moments by $m_j m_k$. Diagonal shrinkage does not guarantee positive definiteness for inconsistent released moments.

**Calibration from score bins.** Freeze $r$, then fit $\widehat{q}(x) = \sigma(a_{\text{cal}} + b_{\text{cal}} r(x))$, where $\sigma(z) = (1+e^{-z})^{-1}$. For compliant bins $B_h$, release counts $N_h = |B_h|$, $K_h = \sum_{i \in B_h} c_i$, and mean scores $\overline{r}_h = N_h^{-1}\sum_{i\in B_h} r(x_i)$, with disclosure protection. The grouped objective is

$$\ell_{\text{bin}}(a_{\text{cal}}, b_{\text{cal}}) = \sum_h \left[K_h z_h - N_h \log(1+e^{z_h})\right], \qquad z_h = a_{\text{cal}} + b_{\text{cal}}\overline{r}_h.$$

Replacing individual scores by their bin mean approximates row-level logistic calibration, even with exact counts, unless scores are constant within bins (or another special equality holds). Rounding adds a separate approximation. If $b_{\text{cal}} > 0$, calibration preserves all score orderings and ties. With $b_{\text{cal}} = 0$ it collapses them, and with $b_{\text{cal}} < 0$ it reverses them. The fitted probability targets $C$, not latent disease.

**Disease and code rankings under class-conditional coding.** Suppose $C \perp X \mid Y$, $a = P(C=1 \mid Y=1) > b = P(C=1 \mid Y=0)$, and both $Y$ and $C$ have nonzero probabilities for each class. Then $q(x) = b + (a-b)\eta(x)$, so true code risks and true disease risks order patients identically. This statement concerns the true risks, not the accuracy of any fitted score. For any fixed score $r$, with arbitrary features, evaluated on independent observations, write $\rho_c = P(Y=1 \mid C=c)$. The established mixture relation [1], including tied scores, is

$$\text{AUC}_C(r) - \tfrac{1}{2} = (\rho_1 - \rho_0)\{\text{AUC}_Y(r) - \tfrac{1}{2}\}. \tag{1}$$

Here $\text{AUC}_Z(r) = P(r(X_Z^+) > r(X_Z^-)) + \frac{1}{2}P(r(X_Z^+) = r(X_Z^-))$, where $X_Z^+, X_Z^-$ are independent draws conditional on $Z=1, Z=0$, respectively, for $Z \in \{Y, C\}$.

*Proof.* Let $H(u,v) = \mathbf{1}\{u > v\} + \frac{1}{2}\mathbf{1}\{u = v\}$ and $A = \text{AUC}_Y(r)$. Since $H(u,v) + H(v,u) = 1$, two independent scores from the same disease class have mean comparison 1/2, including ties. Conditional independence makes each code-class score distribution a mixture of the two disease-class distributions, with positive mixture weight $\rho_c$. Expanding the independent pair gives

$$\begin{aligned}\text{AUC}_C(r) &= \tfrac{1}{2}\{\rho_1\rho_0 + (1-\rho_1)(1-\rho_0)\} + \rho_1(1-\rho_0)A + (1-\rho_1)\rho_0(1-A) \\ &= \tfrac{1}{2} + (\rho_1 - \rho_0)(A - \tfrac{1}{2}).\end{aligned}$$

Finally, writing $\pi_Y = P(Y=1)$ and $p_C = P(C=1)$, Bayes' rule gives $\rho_1 - \rho_0 = \pi_Y(1-\pi_Y)(a-b)/[p_C(1-p_C)] > 0$. This proves the identity and its positive scaling. □

Consequently, on this same population the ordering of fixed models by *population* code AUROC equals their ordering by disease AUROC. Finite-sample AUROCs can rank models differently through sampling error, and the identity does not identify individual disease labels or validate estimated disease probabilities. It is conditional on a frozen fitted rule, with evaluation observations independent of fitting. It does not give the same affine correction for AUPRC or calibration. With no false-positive codes ($b = 0$) and constant positive-case coding probability $a > 0$ (selected completely at random), $\rho_1 = 1$, $\rho_0 = \alpha = (\pi_Y - p_C)/(1 - p_C)$, recovering $\mathrm{AUC}_C = (1 - \alpha)\,\mathrm{AUC}_Y + \alpha/2$ as the positive-unlabeled special case.

**Alternative coding assumptions.** If coding is class-conditional only within group $G = g$, the same results hold within each group with its own coding rates. Different group-specific maps do not ensure pooled patient or model ordering. Alternatively, no false positives and a *known* feature-dependent coding probability $e(x) = P(C = 1 \mid Y = 1, X = x) > 0$ imply $q(x) = e(x)\eta(x)$, hence $\eta(x) = q(x)/e(x)$. We do not know $e(x)$, and HPS group prevalence does not identify it. These coding mechanisms are interpretive assumptions, not empirically established properties of this cohort.

Table S5 illustrates the no-false-positive special case for a coded share of 0.011 and the cross-validated elastic-net AUROC of 0.680. The disease prevalences are hypothetical for the population still uncoded at day 90. They are not estimates from the Household Pulse Survey. Here $\pi_{90} = P(Y = 1)$ and $\alpha_{90} = P(Y = 1 \mid C = 0)$ for that population.

**Table S5: Implied disease AUROC under hypothetical disease prevalence among patients uncoded at day 90.**

| $\pi_{90}$ | $\alpha_{90}$ | Implied disease AUROC |
|---|---|---|
| 0.02 | 0.009 | 0.682 |
| 0.05 | 0.039 | 0.687 |
| 0.10 | 0.090 | 0.698 |
| 0.15 | 0.141 | 0.709 |
| 0.20 | 0.191 | 0.723 |
| 0.30 | 0.292 | 0.754 |

#### S6. Supplementary methods

*Summary-count candidates for the application.* From each feature's rounded $2 \times 2$ table with the outcome (minus the expected rounding excess of 4.5 per cell), naive Bayes uses weights equal to the log odds ratio $w_j$. The empirical-Bayes version replaces $w_j$ by $\tilde{w}_j = w_j \tau^2/(\tau^2 + \mathrm{se}_j^2)$, with $\mathrm{se}_j^2$ the sum of reciprocal cell counts and $\tau^2 = \max\{0, \overline{w^2} - \overline{\mathrm{se}^2}\}$. Linear discriminants use $w = \Sigma^{-1}(\mu_1 - \mu_0)$, where class means come from the label tables and the within-class covariance $\Sigma_W$ from released pair counts (independence values where we suppressed a pair). We shrank $\Sigma_W$ as $(1-\lambda)\Sigma_W + \lambda\,\mathrm{diag}(\Sigma_W)$ for $\lambda \in \{0.1, 0.3, 0.6\}$ and fit each version with and without empirical-Bayes shrinkage of the standardized mean differences.

*Rounding perturbation.* Across 200 replicates that replaced each rounded count independently by a compatible value, refit naive Bayes weights correlated at least 0.95 with the published weights (median 0.97), and the synthetic high-risk patient stayed in the High band, with the central 95% of its predicted probabilities at about 5.5% to 6.3%. We perturbed counts independently, without enforcing joint compatibility. The within-class covariance matrix built from the counts can be slightly indefinite (smallest eigenvalue across replicates −0.0004), which shrinkage toward the diagonal mitigates.

*Laboratory results and vital signs.* We counted a laboratory result in days 0 to 90 as abnormal if it lay outside the recorded reference range in the direction that applies to the test (above the upper bound for most tests, below the lower bound for vitamin D and B12, outside either bound for TSH), if its value concept indicated an abnormal result (for example, high, low, abnormal, positive, detected, or reactive), or, when the bound for that direction was missing, beyond unit-specific thresholds (for example, D-dimer above 0.5 $\mu$g/mL FEU, C-reactive protein above 10 mg/L, ferritin above 300 ng/mL). We read value concepts with negation-aware word rules: a flag word preceded by "not," "non," "no," "none," or "without," or followed by "normal," did not count, so "non-reactive" and "high normal" were not abnormal. Each direction used its own recorded bound, and we matched units on their UCUM codes as well as their names. Vital-sign features were any heart rate above 100 per minute in days 31 to 90, a mean heart rate in days 31 to 90 above 90 per minute or at least 10 per minute above the baseline mean, and any oxygen saturation below 94% in days 0 to 90. Each laboratory test had a result in days 0 to 90 for 1% to 16% of patients, and 16% to 89% of tested participants had at least one result with a recorded upper reference bound. When we added the laboratory-result and vital-sign features to the 102 features of matrix A, the cross-validated AUROC was 0.682 (95% CI 0.643–0.719), against 0.688 (0.651–0.725) without them (a paired difference of −0.006, 95% CI −0.012 to 0.001), and the held-out-site AUROC was 0.624 (0.546–0.715) against 0.618 (0.544–0.722) (a difference of 0.006, 95% CI −0.012 to 0.026).

*Survey reweighting.* HPS reports ages 18–29 and 30–39, and 70–79 and 80 or older, separately. In the primary analysis, we combined them with fixed weights of 0.48 and 0.52, and 0.65 and 0.35. For composition reweighting, we raked HPS person weights among adults who had COVID-19, separately for women and men, to the All of Us cohort's age-band and race-and-ethnicity margins for that sex.

#### S7. Alternative model families

We tuned gradient-boosted trees by an inner three-fold cross-validation over tree depth (2 to 4), learning rate (0.03 or 0.1), and number of trees (200 or 400), nested inside the outer folds so that tuning never saw the evaluation fold. We used scikit-learn's histogram-based implementation. The random forest had 500 trees with at least 20 patients per leaf, and the decision tree had depth 4 with at least 100 patients per leaf. These analyses used the feature matrix, outcome, and evaluation partitions of Table S10.

**Table S6: Alternative model families for a subsequent U09.9 code at day 90.** 95% bootstrap intervals (200 replicates). We pooled the held-out sites (5,760 patients).

| Model | AUROC (95% CI) | AUPRC |
|---|---|---|
| *Five-fold cross-validation* | | |
| random forest | 0.672 (0.642–0.698) | 0.023 |
| decision tree (depth 4) | 0.614 (0.582–0.650) | 0.019 |
| elastic net (M2) | 0.680 (0.651–0.710) | 0.034 |
| tuned boosted trees (scikit-learn, nested tuning) | 0.648 (0.616–0.680) | 0.023 |
| *Held-out sites* | | |
| random forest | 0.665 (0.588–0.734) | 0.025 |
| decision tree (depth 4) | 0.619 (0.538–0.698) | 0.017 |
| elastic net (M2) | 0.618 (0.523–0.713) | 0.025 |
| tuned boosted trees (scikit-learn, nested tuning) | 0.624 (0.524–0.706) | 0.024 |

#### S8. Richer feature representation and additional comparisons

The chosen summary release uses binary features to form $2 \times 2$ tables. Binary encoding is a design choice for this release, and exact NB and LDA summaries can also be defined for other feature types. We compared the binary feature set with an exploratory feature set containing continuous age and event counts, refitting participant-level models on the day-90 landmark cohort with these richer features: continuous age and its square, the number of symptom clusters coded in days 31 to 90, and log counts of visits by class (outpatient, emergency, inpatient) and window, of distinct coding dates for every condition group before and after the index, and of distinct dates of each test, laboratory order, and prescription. Condition codes came from both the condition and observation tables. We fit an elastic net ($\ell_1$ ratio 0.5, $C = 0.05$) on the binary features (reproducing M2), on the rich features, and on both. We also fit an elastic net on the rich features with $C$ chosen by inner three-fold cross-validation from 0.01, 0.05, 0.2, and 1, and scikit-learn histogram gradient boosting on the rich features (300 iterations, learning rate 0.05, 15 leaves, at least 40 patients per leaf). Folds and held-out sites were those of Table S10. At the held-out sites, the released LDA reached an AUROC of 0.643 (main text) and the unselected NB candidate 0.659 (0.576–0.722, Section S10).

**Table S7: Binary and richer features for a subsequent U09.9 code at day 90.** 95% bootstrap intervals (200 replicates). We pooled the held-out sites (5,760 patients).

| Model | Features | AUROC (95% CI) | AUPRC |
|---|---|---|---|
| *Five-fold cross-validation* | | | |
| elastic net, binary features (as M2) | 102 | 0.680 (0.651–0.710) | 0.034 |
| elastic net, rich features | 123 | 0.692 (0.659–0.723) | 0.033 |
| elastic net, binary plus rich features | 225 | 0.705 (0.674–0.737) | 0.042 |
| elastic net, rich features, tuned penalty (C 0.01 or 0.05) | 123 | 0.691 (0.660–0.728) | 0.029 |
| gradient boosting, rich features | 123 | 0.694 (0.666–0.723) | 0.034 |
| *Held-out sites* | | | |
| elastic net, binary features (as M2) | 102 | 0.618 (0.523–0.713) | 0.025 |
| elastic net, rich features | 123 | 0.600 (0.520–0.678) | 0.016 |
| elastic net, binary plus rich features | 225 | 0.617 (0.524–0.702) | 0.022 |
| elastic net, rich features, tuned penalty (C 0.01 or 0.05) | 123 | 0.673 (0.594–0.750) | 0.019 |
| gradient boosting, rich features | 123 | 0.708 (0.647–0.764) | 0.019 |

#### S9. Development and evaluation populations and source grouping

Table S8 compares the development partitions and test sets of the random patient split and the site split. Table S9 describes the source identifiers that define the sites. We defined each patient's site by the source of the index event. Records in the feature tables could come from other sources, so the held-out sites were not fully separated from the development sites.

**Table S8: Development and evaluation populations at day 90.** The random split divides the landmark cohort into an 80% development partition and the 20% patient test. The site split divides it into the 18 held-out sites (the site test) and all other sites (the development partition). Each pair of columns partitions the landmark cohort. Values are n (%). We rounded counts up to a multiple of 10 and computed percentages from the rounded counts. ≤20: 20 or fewer patients. –: we withheld the count so that a reader cannot recover the suppressed count from the other cells (complementary suppression).

| | Random split | | Site split | |
|---|---|---|---|---|
| | Development | Patient test | Held-out sites | Development sites |
| Patients, n | 18,620 | 4,680 | 5,760 | 17,540 |
| First U09.9 code in days 91–455 | 210 (1.1) | 50 (1.1) | 60 (1.0) | 210 (1.2) |
| *Sex* | | | | |
| Female | 11,640 (62.5) | 2,920 (62.4) | 4,000 (69.4) | 10,560 (60.2) |
| Male | 6,830 (36.7) | 1,730 (37.0) | 1,710 (29.7) | 6,850 (39.1) |
| Other/unknown | 160 (0.9) | 40 (0.9) | 60 (1.0) | 140 (0.8) |
| *Age, years* | | | | |
| 18–39 | 2,990 (16.1) | 780 (16.7) | 1,050 (18.2) | 2,720 (15.5) |
| 40–49 | 2,530 (13.6) | 600 (12.8) | 810 (14.1) | 2,330 (13.3) |
| 50–59 | 3,410 (18.3) | 850 (18.2) | 1,060 (18.4) | 3,200 (18.2) |
| 60–69 | 4,730 (25.4) | 1,210 (25.9) | 1,470 (25.5) | 4,470 (25.5) |
| 70+ | 4,990 (26.8) | 1,250 (26.7) | 1,380 (24.0) | 4,850 (27.7) |
| *Race and ethnicity* | | | | |
| Hispanic or Latino | 2,920 (15.7) | 740 (15.8) | 350 (6.1) | 3,310 (18.9) |
| NH Asian | 490 (2.6) | 140 (3.0) | 90 (1.6) | 540 (3.1) |
| NH Black | 2,290 (12.3) | 560 (12.0) | 710 (12.3) | 2,140 (12.2) |
| NH White | 11,410 (61.3) | 2,890 (61.8) | 4,240 (73.6) | 10,060 (57.4) |
| NH other or multiple | 1,000 (5.4) | 250 (5.3) | 240 (4.2) | 1,000 (5.7) |
| Unknown | 540 (2.9) | 130 (2.8) | 150 (2.6) | 520 (3.0) |
| *Chart features by day 90* | | | | |
| Hospitalized around the COVID-19 diagnosis | 2,590 (13.9) | 600 (12.8) | 700 (12.2) | 2,490 (14.2) |
| Emergency visit in days 0 to 30 | 2,830 (15.2) | 680 (14.5) | 930 (16.1) | 2,580 (14.7) |
| 4+ outpatient visits in days −365 to −91 | 14,490 (77.8) | 3,630 (77.6) | 4,600 (79.9) | 13,510 (77.0) |
| 2+ outpatient visits in days 31 to 90 | 11,710 (62.9) | 2,920 (62.4) | 3,470 (60.2) | 11,160 (63.6) |
| Two or more new symptom types | 2,770 (14.9) | 700 (15.0) | 760 (13.2) | 2,710 (15.5) |
| COVID-19 still coded in days 31 to 90 | 1,050 (5.6) | 290 (6.2) | 200 (3.5) | 1,130 (6.4) |
| History of obesity | 5,210 (28.0) | 1,300 (27.8) | 1,500 (26.0) | 5,010 (28.6) |
| History of hypertension | 8,630 (46.3) | 2,090 (44.7) | 2,460 (42.7) | 8,250 (47.0) |
| History of depression | 4,310 (23.1) | 1,050 (22.4) | 1,460 (25.3) | 3,910 (22.3) |

**Table S9: Source identifiers and records from other sources.** Day-90 landmark cohort. We assigned each participant the source identifier of the index event. The participant-mediated EHR sources are PTSC (Participant Technology Systems Center) and TPC (The Participant Center). We counted a patient when they had at least one record of the kind described in days −365 to 90. The lower panel gives the share of each feature table's records in days −365 to 90 that came from a source other than the participant's index-event source (no record in any table lacked a source identifier). We rounded counts up to a multiple of 10 and computed percentages from the rounded counts.

| Measure | Patients, n (%) |
|---|---|
| *Landmark cohort (23,300 patients)* | |
| Index-event source type: EHR site | 21,770 (93.4) |
| Index-event source type: participant-mediated EHR, PTSC | 1,100 (4.7) |
| Index-event source type: participant-mediated EHR, TPC | 440 (1.9) |
| Index-event source identifier missing | 0 (0.0) |
| Index day carries U07.1 records from more than one source | 220 (0.9) |
| Any record in days −365 to 90 from a source other than the index-event source | 12,430 (53.3) |
| Any record in days −365 to 90 with a missing source identifier | 0 (0.0) |
| No record in days −365 to 90 in the feature tables | 0 (0.0) |
| Distinct index-event source identifiers | 54 |
| Distinct source identifiers on feature-table records | 67 |
| *Held-out sites (5,760 patients)* | |
| Any record in days −365 to 90 from a training-site source | 710 (12.3) |
| *Training sites (17,540 patients)* | |
| Any record in days −365 to 90 from a held-out-site source | 100 (0.6) |
| **Records in days −365 to 90 from a source other than the index-event source, by feature table** | **Records, %** |
| Condition | 6.5 |
| Observation | 46.3 |
| Visit | 9.5 |
| Procedure | 9.2 |
| Measurement | 9.5 |
| Drug exposure | 15.0 |

#### S10. Additional prediction and implementation analyses

**Cross-validation and model families.** We evaluated prevalence alone (M0), logistic regression on eight healthcare-use indicators (M1), elastic-net logistic regression (M2), histogram gradient boosting (M3), and bagged positive-unlabeled learning (M4). The elastic net used an $\ell_1$ mixing proportion of 0.5 and inverse penalty parameter 0.05. In each of 25 PU bags, a gradient-boosting classifier contrasted all coded patients with a bootstrap sample of uncoded patients, capped at five times the number of coded patients. The mean of the 25 classifier outputs was a ranking score, because the sampled class proportions do not calibrate it to cohort risk [2]. Five-fold stratified cross-validation used all 23,300 eligible patients. For site evaluation, models were fitted to all development-site patients, without a separate calibration sample. Table S10 reports discrimination, with 200 participant bootstrap replicates and fixed predictions.

**Table S10: Additional model comparisons for subsequent U09.9 at day 90.** AUROC and AUPRC with 95% bootstrap intervals. Held-out sites are pooled.

| Model | AUROC (95% CI) | AUPRC (95% CI) |
|---|---|---|
| *Five-fold cross-validation (23,300 patients)* | | |
| M0 prevalence only | 0.500 | 0.011 |
| M1 utilization (logistic) | 0.574 (0.537–0.610) | 0.015 (0.012–0.019) |
| M2 elastic net | 0.680 (0.643–0.712) | 0.034 (0.024–0.048) |
| M3 gradient boosting | 0.681 (0.650–0.709) | 0.032 (0.022–0.051) |
| M4 bagged PU learning | 0.663 (0.632–0.695) | 0.029 (0.021–0.044) |
| *Held-out sites (18 sites, 5,760 patients)* | | |
| M1 utilization (logistic) | 0.629 (0.545–0.705) | 0.014 (0.009–0.021) |
| M2 elastic net | 0.618 (0.523–0.704) | 0.025 (0.013–0.052) |
| M3 gradient boosting | 0.658 (0.579–0.728) | 0.030 (0.013–0.077) |
| M4 bagged PU learning | 0.672 (0.588–0.744) | 0.028 (0.015–0.060) |

Patients in the top 5% of out-of-fold elastic-net scores had a coded share of 4.1% and included 19% of later-coded patients. Among those later-coded patients, the median time from the landmark to the code was 55 days (interquartile range, 15–109). At the same 5% selection fraction, the symptom-count rule included 15% of later-coded patients and the utilization model included 12%. The elastic net selected 5.4% of women and 4.4% to 7.0% of the racial, ethnic, and age groups shown in Table S2. The decision curve for the coded outcome favored the elastic net over selecting no patients at thresholds of 0.5% to 5% and over selecting all patients at thresholds of 1% to 5% (Figure S2) [3].

**Subsequent symptom and comparison-condition codes.** Among patients without a U09.9 code during follow-up, we compared code occurrence in days 91–455 between the top 10% and bottom 50% of out-of-fold elastic-net scores. The five symptom groups were fatigue, dyspnea, cognitive symptoms, autonomic symptoms, and dizziness. Comparison conditions were fractures, lacerations, cataract, and appendicitis. For new codes, we excluded patients with a prior code for that event and combined comparisons across 12 healthcare-utilization strata using a Mantel–Haenszel estimator. Confidence intervals used 200 participant bootstrap replicates.

Crude ratios for any later code were 2.9–4.0 for the symptom groups and 1.5–2.7 for fractures, lacerations, and cataract (Figure S4B). Adjusted ratios for new codes were 1.7–2.1 for symptoms, 1.9 for fractures, 2.0 for lacerations, and 1.1 for cataract. Appendicitis had too few events for a stable estimate. The score was therefore associated with several forms of subsequent documentation.

These conditions do not establish a formal negative-control design: post-COVID illness could affect injury risk, and healthcare contact affects whether any event is recorded. The comparisons describe associations with later codes rather than validate latent Long COVID labels.

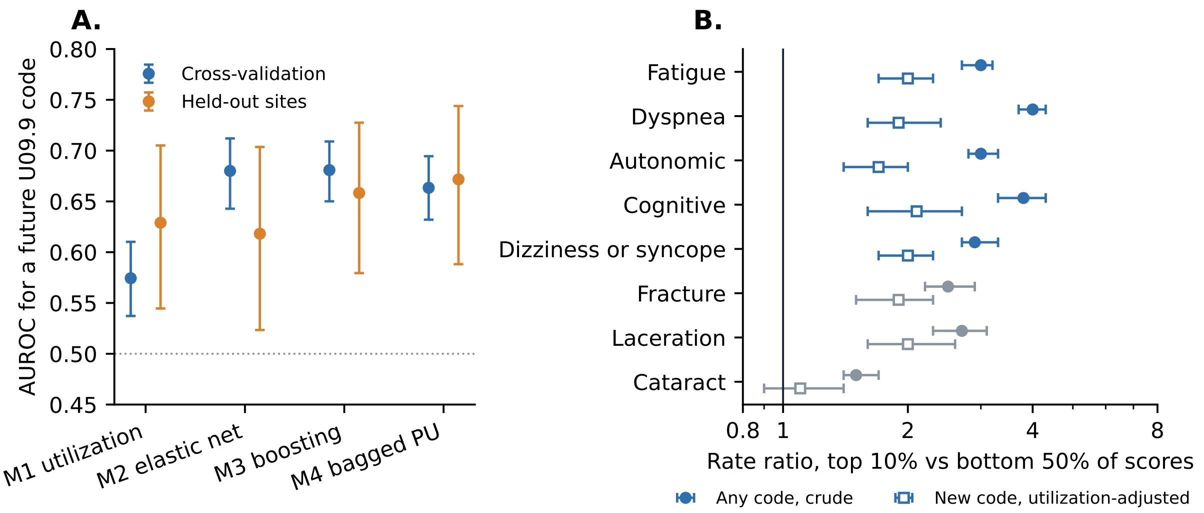


**Figure S4: Additional discrimination and documentation associations.** **A.** AUROC with 95% bootstrap intervals, from five-fold cross-validation and 18 held-out sites. **B.** Among patients without a subsequent U09.9 code, ratios comparing the top 10% with the bottom 50% of elastic-net scores for Long COVID–associated symptom codes and comparison conditions: any code, crude (filled circles), and new codes adjusted for healthcare use (open squares), with 95% bootstrap intervals.

**Landmark and score-adjustment analyses.** We rebuilt eligibility, features, and outcomes at days 30, 60, 90, 120, and 180, fitting a separate elastic net at each landmark [4]. For landmarks before day 90, index-date restrictions ensured that every outcome day followed the introduction of U09.9. AUROC ranged from 0.63 at day 30 to 0.70 at day 180 (Figure S5). Among the same patients eligible at day 180, the day-180 model had an AUROC of 0.68 and the day-30 model 0.62, a paired difference of 0.055 (95% CI, 0.0001–0.113). In a separate exploratory analysis, dividing women's out-of-fold scores by the survey-based women-to-men coding ratio increased their share of the top 5% from 67% to 75% (62% of the cohort). Recall changed from 19% to 23%, a single rounding increment in the coded count, and AUROC changed from 0.688 to 0.684 (Table S3). This adjustment uses a full-cohort coding ratio and does not estimate the day-90 patient's disease probability.

**Application model.** The SMART on FHIR application runs a naive Bayes model with 57 features and empirical-Bayes shrinkage, fitted from counts of the full random development partition (18,620 patients). Of the 102 features, 57 met the four-cell eligibility rule, and 1,596 pair counts were released for them. Table S11 reports eight NB and LDA specifications fitted to these counts and evaluated on the patient test. The application uses NB because each weight is a shrunken log odds ratio that the interface can show as a finding's contribution. Its calibration map came from bins of development scores, each with at least 50 coded and 50 uncoded patients. Figure S6 shows the largest weights. The application computes its inputs from FHIR resources, so its feature definitions differ in places from the research definitions of the released models.

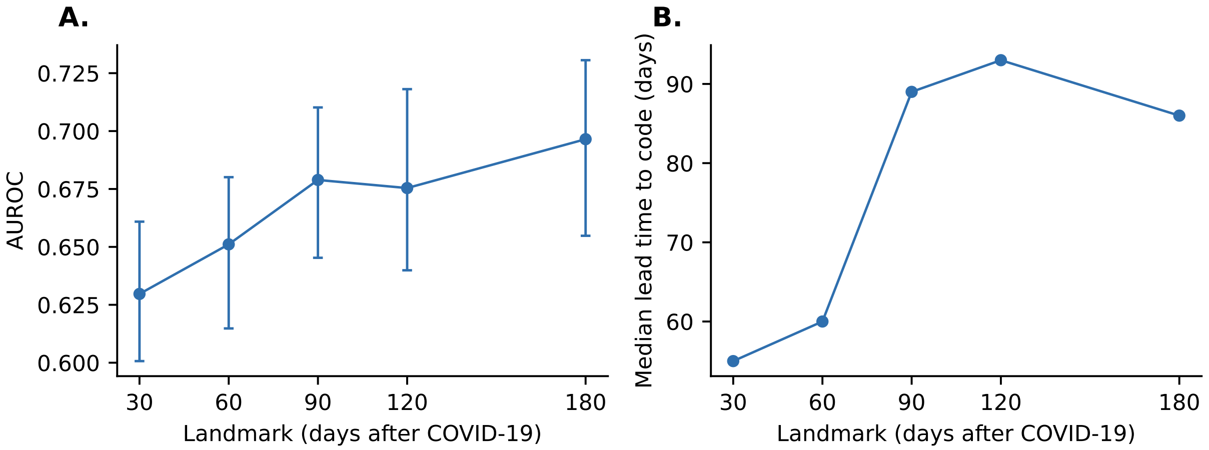


**Figure S5: Landmark-specific prediction tasks.** Each task predicts a first U09.9 code from the day after its landmark to day 455, so eligibility and case composition differ. **A.** AUROC with 95% bootstrap intervals. **B.** Median days from the landmark to the code among later-coded patients in the top 10% of scores.

**Unselected NB candidate in the site partition.** In the site partition, cross-validation within the fitting sample selected LDA over NB by 0.005 in mean AUROC (Table S13). Fitted to the same released counts (12,310 patients, 150 coded), the unselected NB specification had an AUROC of 0.66 (95% CI, 0.58–0.72) at held-out sites, against 0.62 (0.52–0.70) for an elastic net fitted to all development-site patients. The paired AUROC difference was 0.040 (95% CI, −0.028 to 0.109), and AUPRC was 0.018 against 0.025 (difference −0.007, 95% CI, −0.027 to 0.001). With a calibration map from two bins of the same calibration sample, its held-out-site calibration slope was 1.02, intercept −0.27, and observed-to-expected ratio 0.90, without intervals. Selection used only the fitting sample, so this test result did not inform the choice of the released model.

**Healthcare use before documentation.** For later-coded patients, we counted visits and tests from day 91 to the day before their first code and compared rates per 30 days with uncoded patients in the same strata of age, sex, acute hospitalization, and baseline utilization. The median interval was 101 days. Later-coded patients had 6.6 (95% CI, 3.7–9.5) more outpatient visits and 0.4 (0.2–0.7) more emergency visits, together with more echocardiograms, chest CT scans, and pulmonary function tests. At 2024 Medicare allowed amounts, the difference was approximately $900 per patient.

**Table S11: NB and LDA specifications fitted to counts of the full random development partition.** Evaluation on the patient test (4,680 patients, 50 later coded, counts rounded upward to 10).

| Model | AUROC (95% CI) | AUPRC (95% CI) |
|---|---|---|
| *Fit outside the enclave from summary counts (exportable)* | | |
| Naive Bayes | 0.674 (0.592–0.747) | 0.028 (0.016–0.065) |
| Naive Bayes, empirical-Bayes shrinkage (implemented) | 0.674 (0.592–0.747) | 0.028 (0.016–0.066) |
| Linear discriminant, $\lambda = 0.1$ | 0.667 (0.576–0.749) | 0.042 (0.019–0.102) |
| Linear discriminant, $\lambda = 0.1$, empirical Bayes | 0.668 (0.579–0.750) | 0.042 (0.019–0.102) |
| Linear discriminant, $\lambda = 0.3$ | 0.672 (0.584–0.756) | 0.051 (0.019–0.110) |
| Linear discriminant, $\lambda = 0.3$, empirical Bayes | 0.672 (0.582–0.755) | 0.051 (0.019–0.111) |
| Linear discriminant, $\lambda = 0.6$ | 0.679 (0.598–0.756) | 0.036 (0.019–0.091) |
| Linear discriminant, $\lambda = 0.6$, empirical Bayes | 0.680 (0.600–0.755) | 0.035 (0.019–0.088) |
| *Fit on participant rows inside the enclave (not exportable)* | | |
| Elastic net (M2) | 0.680 (0.591–0.757) | 0.089 (0.026–0.172) |
| Bagged PU learning (M4) | 0.673 (0.590–0.751) | 0.040 (0.020–0.096) |

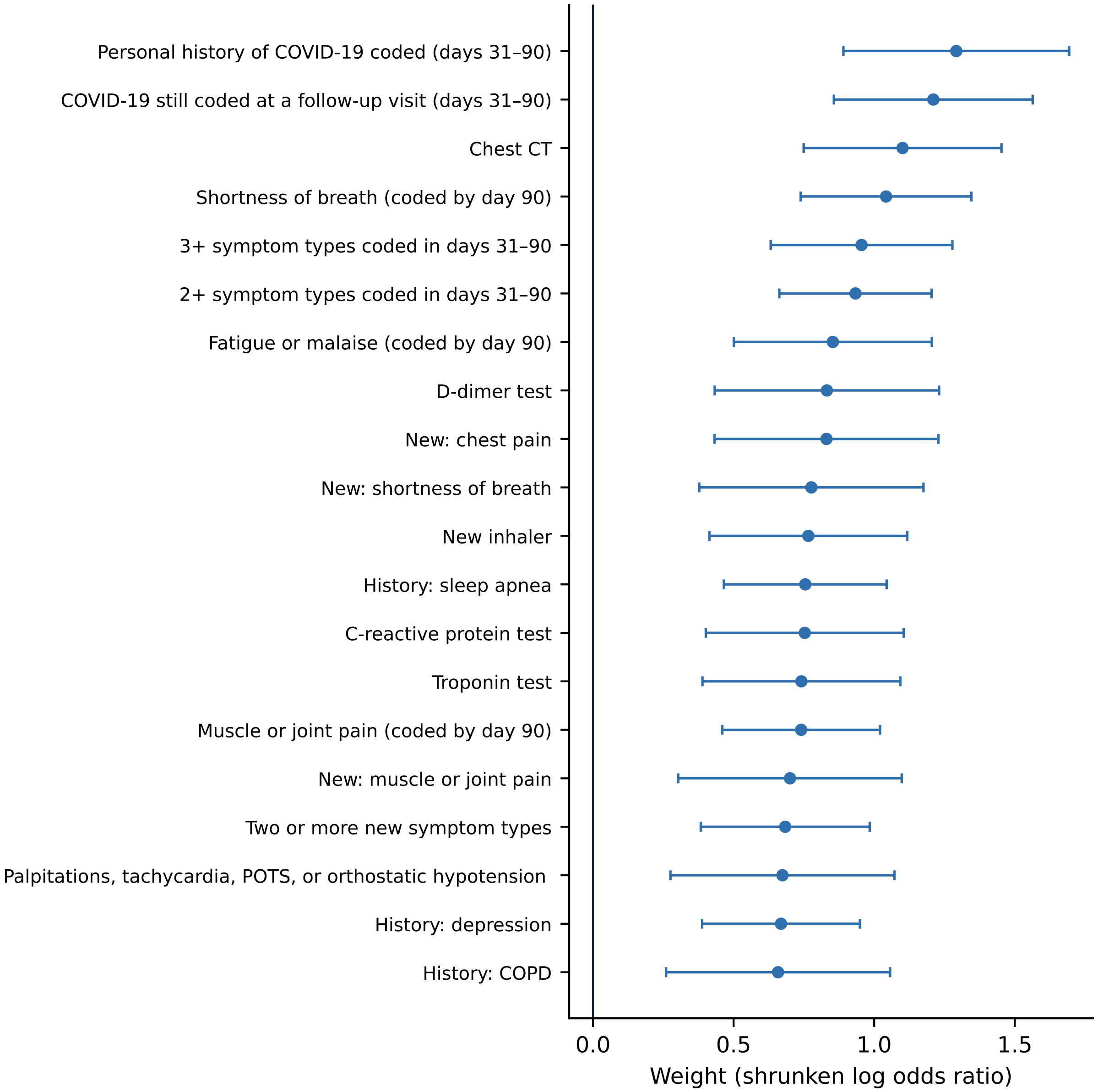


**Figure S6: Largest weights in the 57-feature application model.** Empirical-Bayes shrunken log odds ratios from released counts, with conditional posterior intervals (weight $\pm 1.96\sqrt{k}$ se, where $k$ is the shrinkage factor and the estimated prior variance is treated as known). Each weight contributes to the score when its feature is present.

#### S11. Matched summary-release evaluation

**Partitions and model selection.** The patient partition preserves the random 80% development and 20% test split (seed 20260929). We split its development patients into 70% fitting and 30% calibration samples, stratified by the outcome (seed 20261017). The site partition preserves the 18 held-out sites and the 70% fitting/30% calibration division within development sites (calibration seed 20261003). Table S12 gives protected sample counts. No calibration or test patient contributed to fitting coefficients or selecting the aggregate estimator. These test partitions had also been used in the additional analyses in Section S10.

**Table S12: Samples in the matched evaluation.** Counts are rounded upward to 10 separately, so displayed role counts need not sum to the rounded cohort total.

| Partition | Role | Patients | Coded patients |
|---|---|---|---|
| Random patient test | Fit | 13,040 | 150 |
| | Calibration | 5,590 | 70 |
| | Test | 4,680 | 50 |
| Held-out-site test | Fit | 12,310 | 150 |
| | Calibration | 5,240 | 60 |
| | Test | 5,760 | 60 |

Within each fitting sample, three-fold stratified cross-validation (seed 20261017) compared NB with empirical-Bayes shrinkage and LDA with empirical-Bayes shrinkage and $\lambda \in \{0.1, 0.3, 0.6\}$. In each fold, feature eligibility, count protection, reconstruction, and empirical-Bayes estimation used only that fold's training patients. Mean AUROC selected the specification outside Workbench using released fold metrics. Ties would be broken in the order NB, LDA with $\lambda = 0.1$, 0.3, and 0.6. Table S13 reports the selection metrics. The selected specification was then fitted from summaries of the complete fitting sample. The all-feature and retained-feature exact-count comparators used that same family and shrinkage setting, refitting their empirical-Bayes variances on their respective features. They were not independently tuned to test data.

**Table S13: Fit-only cross-validation for aggregate-model selection.** Unweighted means across three validation folds. EB: empirical-Bayes shrinkage. Selection used AUROC, separately in each partition.

| | Patient partition | | Site partition | |
|---|---|---|---|---|
| Candidate | Mean AUROC | Mean AUPRC | Mean AUROC | Mean AUPRC |
| Naive Bayes, EB | 0.631 | 0.020 | 0.657 | 0.029 |
| LDA, EB, $\lambda = 0.1$ | 0.597 | 0.018 | 0.650 | 0.028 |
| LDA, EB, $\lambda = 0.3$ | 0.607 | 0.018 | 0.655 | 0.029 |
| LDA, EB, $\lambda = 0.6$ | 0.621 | 0.019 | 0.661 | 0.029 |

**Representation and numerical conventions.** The binary matrix contains the 102 indicators in Table S1. The 123 rich features use the same observation windows and code sources, retaining continuous age, age squared, and log event counts in place of binary thresholds (Section S8). Both elastic nets standardize their input columns using fitting-sample means and variances, use the same penalty settings, and use a fixed random seed of 20260928. Thus their contrast includes the chosen representation and the resulting penalized fit.

For the exact-count NB comparator, a zero feature-by-class cell is replaced by 0.5 when calculating its log odds ratio and variance, and positive cells are unchanged. Constant features have zero coefficients. Eligible released features have all four cells greater than 20 before rounding, so this zero-cell convention is not needed for their released NB fit. For LDA, the within-class covariance uses divisor $n$, as derived in Section S5. Standardizing the mean differences uses diagonal variances bounded below by $10^{-9}$. After diagonal shrinkage, the same stabilization rule applies to exact and released covariances. For the resulting matrix $S$, set

$$\epsilon = \max\{10^{-9}, 10^{-8} \max_j D^+_{jj}\}, \qquad \delta = \max\{0, \epsilon - \lambda_{\min}(S)\},$$

where $D^+_{jj} = \max\{D_{jj}, 10^{-9}\}$. The fit solves the linear system using $S + \delta I$. This makes the numerical treatment explicit when rounded counts yield inconsistent moments.

The released fitting summaries contain protected class counts, four-cell feature-by-class tables, feature margins, and retained-feature pair counts. Subtracting 4.5 from positive rounded counts centers their ten compatible integer values but does not identify the original counts. Released margins and pair counts need not agree exactly with the four-cell tables. Unavailable pair moments use the product of reconstructed margins. Both final released fits had complete retained-feature pair tables.

**Calibration and uncertainty.** After the score was fitted outside Workbench, calibration patients were sorted by that fixed score. Adjacent patients were merged until each bin contained at least 21 coded and 21 uncoded patients and at least one twelfth of the calibration sample. A final incomplete bin was merged with its predecessor. The release contained mean scores rounded to four decimal places and protected patient and coded-patient counts. Each partition yielded two informative bins. With two bins, the two-parameter map reproduces both grouped event rates, so the calibration sample offers no check of fit. The test-set estimates below provide that check. The grouped logistic likelihood in Section S5 was fitted outside Workbench with a nonnegative slope, using counts minus 4.5. Both fitted slopes were positive. The complete probability map was returned unchanged for test evaluation, without interpolation or test-set recalibration.

Calibration in the large is the intercept from logistic regression with the artifact's predicted log odds as an offset. Calibration slope comes from a logistic regression with a free intercept and the predicted log odds as its predictor. Both are diagnostic test-set estimates, not changes to the released model. Table S14 gives these estimates and descriptive Brier scores with central 95% percentile intervals from 500 ordinary participant bootstrap samples (seed 20261017). Outcome counts vary in these samples. AUROC and AUPRC use the paired outcome-stratified bootstrap described in the main text. All intervals condition on the already fitted models and chosen settings and exclude uncertainty from refitting or a new sample of sites.

**Artifact identity and reconstruction.** The public package contains separate patient-partition and site-partition instances. The site-partition instance, fitted without the held-out sites, is the focal released model. It records the feature definitions, score coefficients, calibration parameters, selected specification, and hashes of the aggregate inputs and source code. The reference scorer requires explicit binary values for every fitted feature and treats a missing input as an error, not as evidence of absence. Synthetic feature dictionaries supply expected scores and probabilities. Workbench checks compare the imported score with internal reconstruction from protected counts, and compare the complete artifact with the portable reference scorer on every test patient. Only aggregate numerical errors leave Workbench. These checks establish the computational identity of the evaluated artifact. Section S10 describes the separate FHIR implementation.

**Table S14: Test performance of the complete externally calibrated artifacts.** Parentheses give 95% CIs from 500 bootstrap replicates. Calibration and Brier intervals use ordinary participant resampling, and discrimination intervals use outcome-stratified resampling.

| Measure | Random patient test | Held-out-site test |
|---|---|---|
| Calibration intercept in the large | -0.12 (-0.44, 0.15) | -0.29 (-0.58, -0.02) |
| Calibration slope | 0.89 (0.51, 1.19) | 0.83 (0.40, 1.18) |
| Brier score | 0.0105 (0.0077, 0.0136) | 0.0088 (0.0066, 0.0115) |
| AUROC | 0.679 (0.603, 0.753) | 0.643 (0.553, 0.719) |
| AUPRC | 0.030 (0.019, 0.062) | 0.021 (0.012, 0.055) |

**Table S15: Paired differences between matched models.** Each row subtracts the second model from the first, so a negative value means the first, more restricted model performed worse. The first row equals the sum of the next three. 95% CIs from the same 500 paired, outcome-stratified participant bootstrap replicates. The main-text figure plots these values.

| Comparison | AUROC difference (95% CI) | AUPRC difference (95% CI) |
|---|---|---|
| *Random patient test* | | |
| Released counts minus binary elastic net | -0.013 (-0.070, +0.049) | -0.062 (-0.122, -0.003) |
| Exact counts, all features, minus binary elastic net | -0.008 (-0.063, +0.046) | -0.060 (-0.117, -0.003) |
| Retained minus all features (exact counts) | -0.006 (-0.024, +0.013) | -0.001 (-0.032, +0.015) |
| Released minus exact counts (retained features) | +0.001 (-0.006, +0.006) | -0.001 (-0.007, +0.005) |
| Rich minus binary elastic net | -0.004 (-0.084, +0.065) | -0.062 (-0.117, -0.008) |
| *Held-out-site test* | | |
| Released counts minus binary elastic net | +0.049 (+0.001, +0.104) | -0.001 (-0.017, +0.017) |
| Exact counts, all features, minus binary elastic net | +0.026 (-0.002, +0.050) | +0.003 (-0.008, +0.032) |
| Retained minus all features (exact counts) | +0.023 (-0.013, +0.064) | -0.003 (-0.024, +0.005) |
| Released minus exact counts (retained features) | 0.000 (-0.014, +0.013) | 0.000 (-0.009, +0.004) |
| Rich minus binary elastic net | +0.049 (-0.036, +0.128) | -0.006 (-0.030, +0.002) |

### References


[1] Jain S, White M, and Radivojac P. Recovering true classifier performance in positive-unlabeled learning. *Proc AAAI Conf Artif Intell*, 31(1), 2017. doi: 10.1609/aaai.v31i1.10937.

[2] Bekker J and Davis J. Learning from positive and unlabeled data: a survey. *Mach Learn*, 109 (4):719–760, 2020. doi: 10.1007/s10994-020-05877-5.

[3] Vickers AJ and Elkin EB. Decision curve analysis: A novel method for evaluating prediction models. *Med Decis Making*, 26(6):565–574, 2006. doi: 10.1177/0272989x06295361.

[4] van Houwelingen HC. Dynamic prediction by landmarking in event history analysis. *Scand J Stat*, 34(1):70–85, 2007. doi: 10.1111/j.1467-9469.2006.00529.x.
